# Accounting for uncertainty in participant age in serocatalytic models

**DOI:** 10.64898/2026.03.14.26346885

**Authors:** Junjie Chen, Teresa Lambe, Everlyn Kamau, Christl Donnelly, Ben Lambert, Sumali Bajaj

## Abstract

Serological surveys measure the presence of antibodies in a population to infer past exposure to an infectious pathogen. If study participants’ ages are known, serocatalytic models can be used to retrace the historical transmission strength of a pathogen within that population, quantified by the force of infection (FOI). These models rely on age information as a key variable since infection risks are interpreted in relation to how long individuals have been at risk. However, due to data constraints, participants’ ages may be provided only within “age bins”. A common approach is then to assign individuals’ ages to midpoints of their respective age bins, ignoring uncertainty in this quantity. In this study, we quantify the bias introduced by this midpoint approach and develop a Bayesian framework that explicitly accounts for uncertainty in age. By comparing inference under constant, age-dependent, and time-dependent FOI scenarios, we show that the proposed binned model yields more reliable FOI estimates without sacrificing computational complexity, whereas the midpoint approach can underestimate FOI under constant transmission and introduce further biases as age bin width increases. These improvements support the interpretation of serological data and inform public health decisions, such as estimating disease burden and identifying targeted vaccination groups.

## 1 Introduction

Serocatalytic models are powerful tools for inferring historical transmission dynamics of a pathogen using age-stratified serological data. For example, assuming no age-related risk of infection and loss of detectable antibodies (known as *seroreversion*), if individuals under 20 years-old have no antibodies, whereas those above 20 do, this pattern suggests that transmission of that pathogen stopped 20 years ago. Such historical estimates are particularly useful for estimating epidemiological parameters, informing vaccine trial design [1], and inferring infection levels when surveillance is incomplete [2, 3, 4]. Kamau et al. [5] offer an introductory guide to these models and their practical applications.

Unlike transmission models, which describe the progression of infection or predict future outbreaks, serocatalytic models work retrospectively. Their aim is not to forecast, but to reconstruct past transmission based on individuals’ serological results. These results, also known as an individual’s *serostatus*, are typically categorized into binary outcomes: an individual is *seropositive* if their antibody level exceeds a threshold, and *seronegative* otherwise. The primary metric used to quantify past transmission is the *force of infection* (FOI), defined as the average rate at which individuals become infected per unit time. Compared with transmission dynamic models, serocatalytic models typically require fewer assumptions but can still provide insights to support estimation of the key transmission parameters [5].

To derive the serocatalytic model, we follow the notation used by Kamau et al. [5], considering a population with two compartments: *S*^*b*^(*t*), the proportion of individuals born at time *b* who are seronegative at time *t*, and *X*^*b*^(*t*), the proportion born at time *b* who are infected or have recovered from infection and have detectable antibodies against the pathogen at time *t*. Assuming negligible disease-induced mortality, *S*^*b*^(*t*) + *X*^*b*^(*t*) = 1 for all *t*. In the absence of maternal antibodies, all individuals are assumed seronegative at birth, i.e., *S*^*b*^(*b*) = 1.

The proportion of individuals who are seropositive is referred to as the *seroprevalence* or *seropositivity*. To illustrate the basic idea, consider an idealized setting where individuals’ exact ages are known and the FOI is constant, 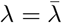. The probability that an individual of age *a* is seropositive can be written as:

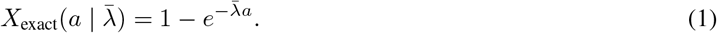

A derivation of Eq.1 is provided in §S1.1. Intuitively, under a constant FOI, the probability of remaining uninfected declines exponentially with age, resulting in a steadily increasing seropositivity that ultimately plateaus at 1.

In reality, the FOI is often not constant. It may be age-dependent, as infection risk can differ across age groups. For example, the risk of chickenpox infection peaks in early childhood [6, 7], whereas HIV risk typically increases during adolescence [7]. FOI can also vary over calendar time due to seasonality, outbreaks, or public health interventions. For example, influenza transmission generally peaks in winter in temperate climates [8]. Derivations of the age- and time-dependent seropositivity expressions are provided in [5].

These different FOI patterns give rise to distinct seropositivity profiles observed from serosurveys, as is illustrated in Figure 1. The top arrow in Figure 1 indicates the forward process that translates FOI into the age-specific seropositivity: the higher the FOI, the faster people become seropositive. In practice, though, the FOI is unobserved. The goal of serocatalytic modeling, represented by the bottom arrow, is to work in the inverse direction: estimating the FOI that gives rise to the observed age-stratified serological data.

**Figure 1:**
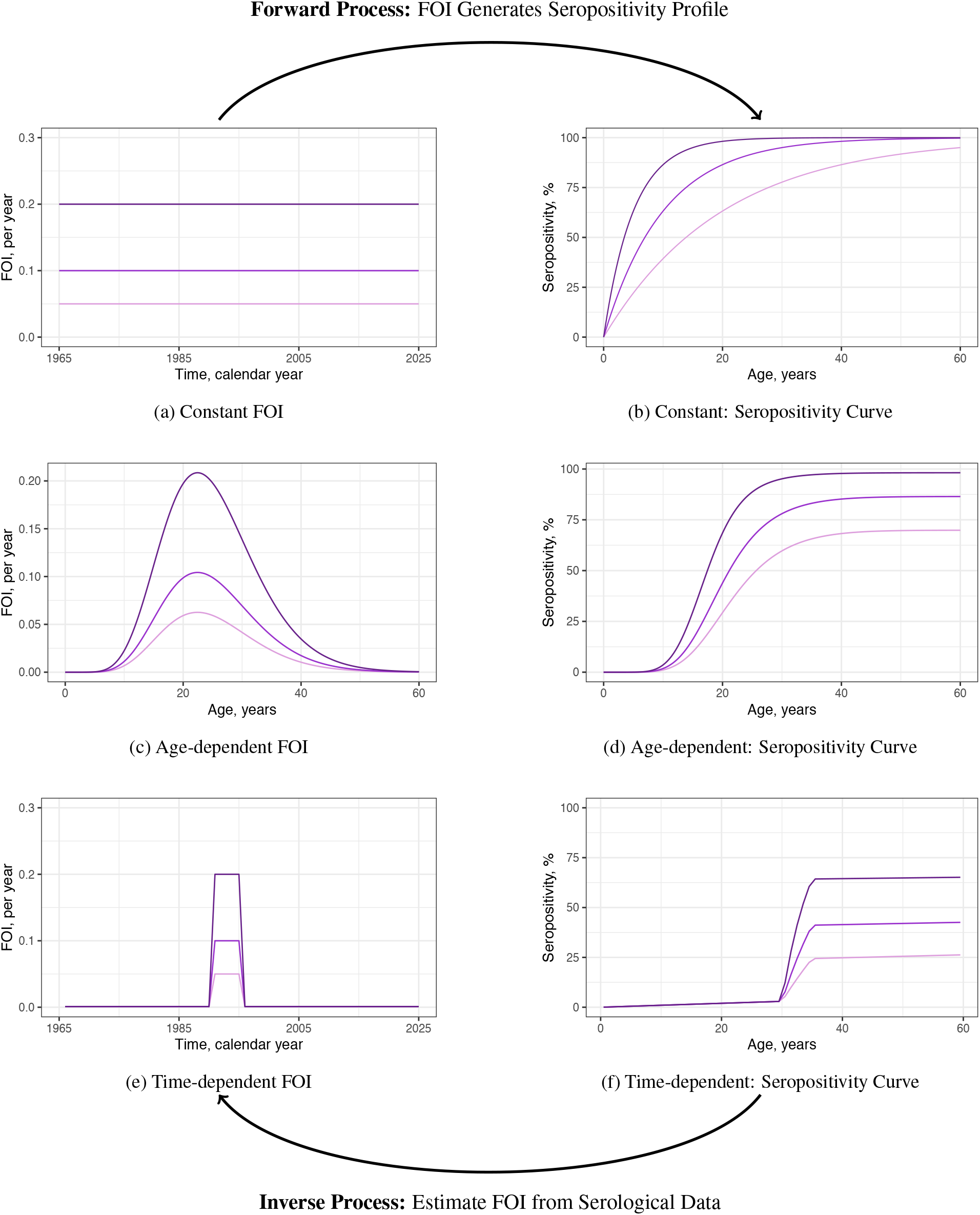
Relationship between the FOI and seropositivity. In each row, the left panel shows the FOI pattern and the right panel shows the resulting seropositivity curve for a serosurvey done in 2025. The top arrow represents the forward process, where different FOI patterns give rise to distinct age-specific seroprevalence profiles. The bottom arrow illustrates the inverse process, where serocatalytic models infer the underlying FOI from observed serological data. Panels (a–b): constant FOI; (c–d): age-dependent FOI following a modal pattern; (e–f): time-dependent FOI, assuming the survey year is 2025.

A key challenge in applying serocatalytic models lies in how age data are recorded in serological surveys. Due to privacy or reporting reasons, age information may only be provided in age groups (or *age bins*) rather than exact values. A recent global review of 66 dengue seroprevalence studies [9] highlights the prevalence of this practice and the need to re-evaluate how such data are used in FOI estimation. A common approach is then to assign each age bin its midpoint as the representative age [10, 11]. While straightforward, this midpoint approximation fails to capture the uncertainty in individuals’ ages within the bin.

In this study, we quantify the bias and the anticipated changes in uncertainty introduced into FOI estimation when ages are reported in bins and compare these results with estimates obtained from our proposed model, which explicitly incorporates uncertainty in age. In particular, we consider three transmission scenarios: constant, age- and time-dependent FOI, and assess model performance using the three types of age data: exact, midpoint, and binned for each scenario. The distinctions between these age types are summarized in Table 1 and explained in the following section. By comparing results across these combinations, we demonstrate the importance of properly accounting for uncertainty in study participants’ age in serocatalytic inference.

**Table 1:** Inputs used in serocatalytic models. The table illustrates how age may be recorded in individual-level serological datasets (exact, binned, or midpoint), together with the individual’s binary serostatus (1 seropositive; 0 seronegative).

| Exact age (years) | Binned age (years) | Midpoint age (years) | Serostatus |
| --- | --- | --- | --- |
| 8.43 | 0–10 | 5 | 0 |
| 12.54 | 10–20 | 15 | 0 |
| 24.56 | 20–30 | 25 | 1 |
| 31.12 | 30–40 | 35 | 0 |
| 45.65 | 40–50 | 45 | 1 |
| 57.42 | 50–60 | 55 | 1 |
| 63.77 | 60–70 | 65 | 1 |

## 2 Methods

Serocatalytic models require two key inputs: serostatus and age, as shown in Table 1. The last column shows each individual’s serostatus *Y*_*i*_, a binary variable indicating whether individual *i* is seropositive (*Y*_*i*_ = 1) or seronegative (*Y*_*i*_ = 0). When exact ages are available, serostatus follows a Bernoulli distribution, with the probability of being seropositive depending on the individual’s age and the FOI they have experienced.

The first three columns in Table 1 illustrate the common formats to record ages in serosurveys. *Exact ages* are rounded to two decimal places for illustration. In the simulated datasets and corresponding inference, they are continuous quantities with the default precision in R. In practice, however, ages are often reported only to the nearest integer year. Although integer ages can be viewed as age groups with a bin width of one year, the additional FOI accumulated within a single year is typically small relative to the overall cumulative risk. We therefore treat integer ages as exact ages in real-world datasets. The second column shows an example of individuals grouped into 10-year age bins *(binned ages)*. The third column presents the midpoint of each age bin, calculated as the average of its lower and upper bounds *(midpoint ages)*.

All models require serostatus as a common input and are implemented within a Bayesian framework (implementation details provided in §S2.1). When exact ages are available, they are used directly in the likelihood under the *exact model*. When only age bins are available, using midpoint ages as substitutes for exact ages is referred to as the *midpoint model*; for the model developed in this study, which explicitly accounts for uncertainty in individuals’ ages within each bin, we refer to it as the *binned model*. The terminologies and color scheme shown in Table 1 are used consistently throughout the paper.

Throughout this paper, we assume there is no seroreversion, such that individuals remain seropositive for life once infected, and that an individual’s serostatus is a perfect measure of their true serostatus. It is straightforward to adapt our method to account for imperfect tests.

### 2.1 Constant FOI Model

We begin with the simplest situation where transmission is endemic and static, i.e., infection risk does not vary by age or time.

#### 2.1.1 Exact Model

When FOI is constant 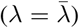 and the exact ages are known, the probability of being seropositive for an individual *i* at age *a*_*i*_ is: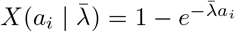, as described by Eq.1. Let *n* denote the total number of individuals in the dataset. Assuming that individuals’ serostatus are conditionally independent given 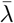, the posterior distribution of the FOI over all *n* serostatus outcomes **Y** = (*Y*_1_, …, *Y*_*n*_) and corresponding ages **a** = (*a*_1_, …, *a*_*n*_) becomes:

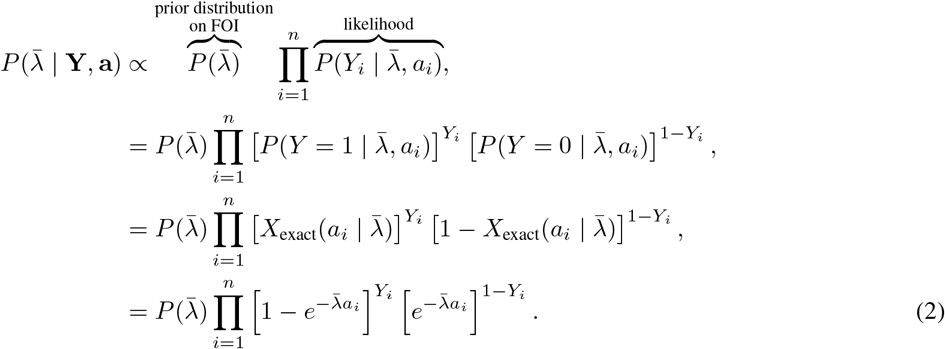

#### 2.1.2 Midpoint Model

The midpoint model also uses Eq.2, treating individuals’ ages as the midpoint of their respective age bins, i.e., 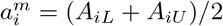 where *A*_*iL*_ and *A*_*iU*_ are the integer lower and upper bounds of the age bin. While this allows midpoint ages to be substituted directly into the exact model likelihood, it ignores the uncertainty in participants’ ages within each bin.

#### 2.1.3 Binned Model

The binned model incorporates uncertainty in ages using the Bayes’ theorem, as in Eq.3. For an individual *i* with observed serostatus *Y*_*i*_ and the corresponding age bin [*A*_*iL*_, *A*_*iU*_ ), the joint posterior distribution of 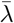 and the unobserved age *a*_*i*_ is:

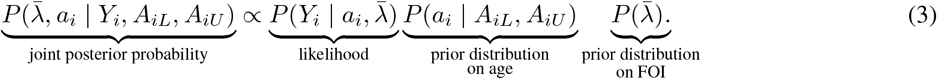

We assume that ages are uniformly distributed within the bin [*A*_*iL*_, *A*_*iU*_ ), which is an assumption we carry throughout this paper. Then, we integrate over all possible ages within each bin to get the marginal posterior distribution of 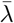 :

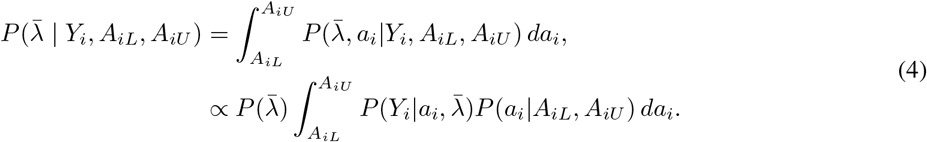

Extending this to all individuals, the overall marginal posterior becomes:

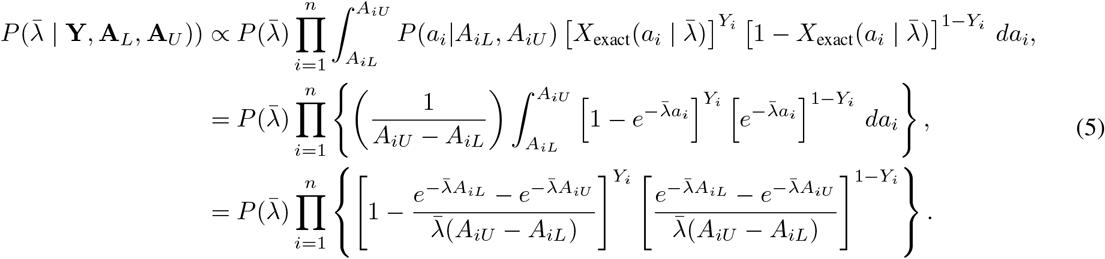

#### 2.1.4 Asymptotic Behavior

As the age bin shrinks to the exact age, *A*_*iL*_ *→ A*_*iU*_ *→ a*_*i*_, and we have:

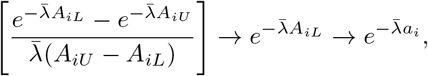

the expression collapses to what we derived when exact ages are known (Eq.2).

### 2.2 Age-dependent FOI Model

When FOI varies with age (*λ*_*a*_), the cumulative risk of infection by age *a*_*i*_, assuming no maternal antibodies, is 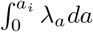. When exact ages are known, seropositivity can be expressed as:

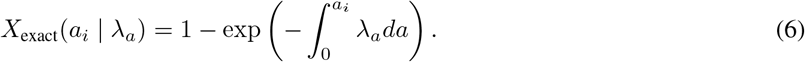

If only the age bin [*A*_*iL*_, *A*_*iU*_ ) is known, we average over all possible ages within the bin:

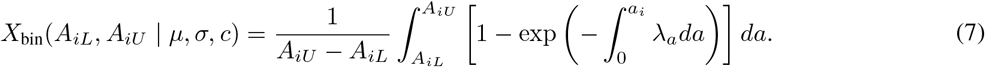

The nested integral in Eq.7 makes further analytical simplification difficult as no closed-form solution exists for the general case. To address this, we explore two modeling approaches based on different assumptions of *λ*_*a*_: (i) a stepwise piecewise-constant function over pre-defined age intervals, and (ii) a continuous parametric function based on the gamma distribution probability density function (PDF).

The piecewise-constant assumption is widely used for its simplicity and flexibility [12, 5, 13, 14], and is described in detail in §S2.2. It assumes FOI is constant within defined age intervals and changes only at fixed cutoffs. However, when applied to age-binned seroprevalence data, evaluating the likelihood becomes complex, which requires additional computational techniques to remain tractable. The corresponding methods and results under the piecewise-constant assumption are presented in §S2.3.

As such, we primarily focus on the second approach, where the FOI is parametrized by a gamma distribution PDF, which provides a smooth and non-negative representation of infection risk across age, while avoiding nested integrals and capturing the unimodal patterns observed in many diseases. Childhood diseases such as measles often show high incidence below 5 years [15, 16], though high overall FOI may also contribute to this pattern. Adult-onset infections, such as HIV, typically peak between 25–35 years [17], whereas herpes zoster is more common in groups over 50 years old [18, 19].

#### 2.2.1 Exact Model

The age-dependent FOI following a gamma distribution PDF is modeled as:

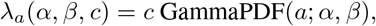

where *c* is a scaling constant that determines the overall transmission intensity, and GammaPDF(*a*; *α, β*) denotes the gamma PDF evaluated at age *a* with shape *α* and rate *β*. For interpretability, we re-parametrize it in terms of mean *µ* and standard deviation *σ*, using the relationships:

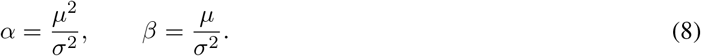

When exact ages are available, the probability of an individual *i* being seropositive by age *a*_*i*_ is:

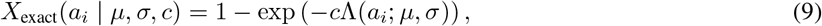

where Λ(*a*_*i*_; *µ, σ*) denotes the gamma cumulative distribution function (CDF), which is the cumulative FOI up to age *a*_*i*_.

Following the same derivation from Eq.2. The resulting posterior distribution of the age-dependent FOI is:

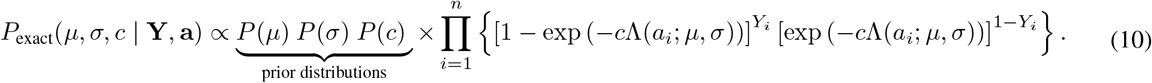

#### 2.2.2 Midpoint Model

Similarly to the constant FOI case, the midpoint model substitutes the midpoint age 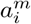 for the exact age *a*_*i*_ in Eq.10, and we now estimate the gamma parameters (*µ, σ, c*) that describe the age-dependent FOI.

#### 2.2.3 Binned Model

When exact ages are unavailable, we incorporate uncertainty in age using Bayes’ theorem, as in the constant FOI case (Eq.3). For an individual *i*, the joint posterior over the model parameters and unobserved age *a*_*i*_ is:

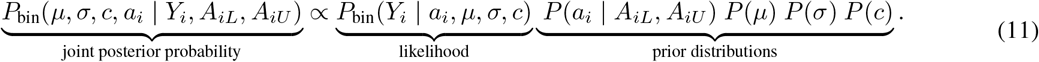

Like in Eq.4, we marginalize over all possible ages within the bin to derive the marginal posterior distribution over *µ, σ* and *c*.

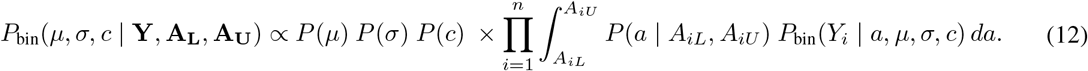

An individuals’ serostatus follows still a Bernoulli distribution, where the probability of being seropositive is given by Eq.9. Substituting this into the likelihood gives:

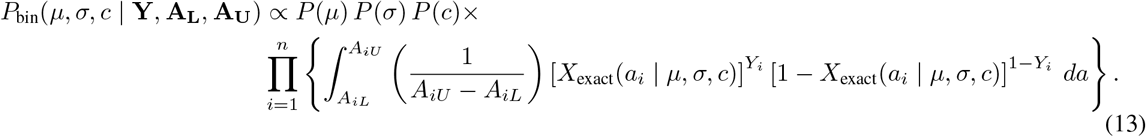

The integrals in Eq.13 have no analytical solution and here we evaluate them numerically using numerical quadrature, as implemented by Stan’s integrate_1d function [20]. This approach approximates the integral over each age bin by discretizing it into a sum, with the numerical accuracy controlled by a relative tolerance, which we set to 1%.

We show in §S2.4.1 that the binned model converges to the exact model as the age bin width tends to zero.

### 2.3 Time-dependent FOI Model

Unlike the age-dependent FOI model, which shows higher infection risk in a specific age cohort, time-dependent patterns are typically irregular. Outbreaks can occur sporadically, sometimes with multiple waves, and seasonality can introduce strong cyclical patterns, especially for vector-borne or respiratory infections [21]. External factors, such as vector control strategies, vaccination, and changing social behavior, can significantly affect transmission dynamics [22]. These features make the smooth and unimodal structure of a model function poorly suited to capture the temporal heterogeneity in FOI. Therefore, we assume FOI is piecewise-constant for the time-dependent model. In this model, we fix a survey time *T*, with individuals born on different dates *b*_*i*_, representing a cross-sectional population observed at time *T* .

#### 2.3.1 Binned Model

For an individual *i* born on date *b*_*i*_ and surveyed at time *T*, seropositivity given by Eq.14. The derivation of this equation and the notation used in it are summarized in Table 2. The boxes under the equation corresponds to the visual breakdown of the binned model presented in Figure S3, and the explanations of each breakdown component can be found in Table S3.

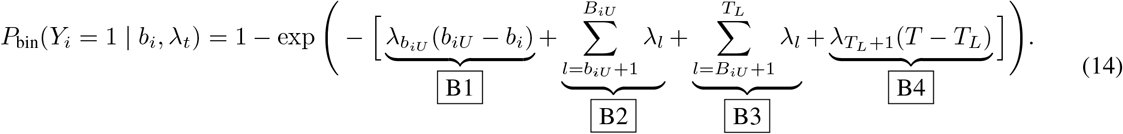

**Table 2:** Notation used in the time-dependent FOI model. Exact ages and numeric dates are rounded to two decimal places for illustration. In the simulations and model inference, ages and dates are continuous variables.

| Notation | Description | Example |
| --- | --- | --- |
| $a_i$ | Exact age of an individual $i$ at the time of conducting the serosurvey. | An individual of age 6.73, i.e., $a_i = 6.73$. |
| $T$ | Fixed date up to which FOI is accumulated (in numeric form<sup>a</sup>). | Current date or sampled date: $T = 08/03/2025$ (2025.18 in numeric form). |
| $T_L$ | Integer year of the fixed date $T$. | For $T = 2025.18$, $T_L = 2025$. |
| $b_i$ | Birth date in numeric form, defined as $b_i = T - a_i$. | If $a_i = 6.73$ and $T = 2025.18$, then $b_i = 2018.45$ (15/06/2018 in dd/mm/yyyy form). |
| $b_{iU}$ | Integer upper boundary of the birth year, i.e., individuals were born before that year. | If $b_i = 2018.45$, then $b_{iU} = 2019$. |
| $B_{iL}$ | Lower boundary of the birth interval, $B_{iL} = T_L - A_{iU}$. | If age bin is $[5, 10)$, then $B_{iL} = 2025 - 10 = 2015$. |
| $B_{iU}$ | Upper boundary of the birth interval, $B_{iU} = T_L - A_{iL}$. | If age bin is $[5, 10)$, then $B_{iU} = 2025 - 5 = 2020$. |
| $\lambda_t$ | Time-dependent FOI for the interval $[t-1, t)$. | For $t = 2026$, $\lambda_{2026}$ represents the FOI for the interval $[2025, 2026)$. |

<sup>a</sup> Numeric dates are computed from a date stamp by adding the fraction of the year elapsed, where 365.25 represents the average number of days per year accounting for leap years: numeric date = year + (day of year - 1)/365.25.

For readers interested in the detailed derivations of the time-dependent FOI model, we refer to §S2.5. In brief, when only binned ages are available, the exact birth date *b*_*i*_ is unknown. By assuming a uniform distribution of *b*_*i*_ within each interval, we marginalize *P*_bin_(*Y*_*i*_ = 1 *b*_*i*_, λ_*t*_) over the birth interval [*B*_*iL*_, *B*_*iU*_ ). This leads to the closed-form expression for the marginal probability of being seropositive:

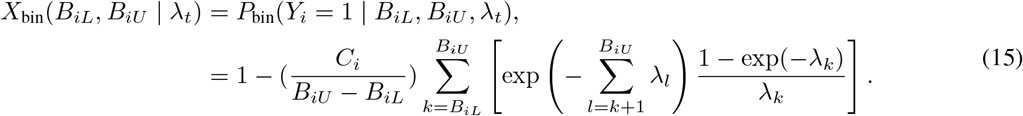

Conditional on λ_*t*_, each serostatus outcome follows a Bernoulli distribution with success probability *X*_bin_(*B*_*iL*_, *B*_*iU*_ λ_*t*_). We define *L*_*i*_(λ_*t*_) as the probability of observing individual *i*’s serostatus under the time-dependent FOI, after accounting for uncertainty in their exact birth date.

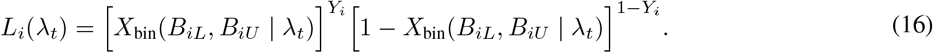

Assuming conditional independence across individuals given λ_*t*_, the posterior distribution of the time-dependent FOI is given by:

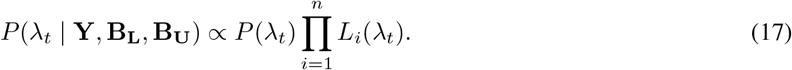

#### 2.3.2 Exact Model

When exact ages are available, individuals’ birth dates *b*_*i*_ are known and there is no need to marginalize over birth intervals. The seropositivity under the exact model is:

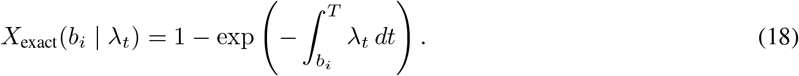

Under the piecewise-constant FOI assumption, the integral in Eq. 18 can be decomposed into contributions from the time intervals in which the FOI is constant:

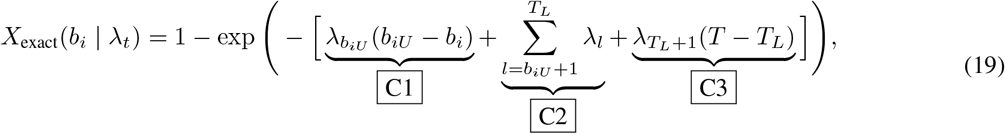

where the three terms correspond to the exact-model components illustrated in Figure S3.

We also confirm the asymptotic behavior of the binned model as the birth interval shrinks to the exact birth year in §S2.5.2 numerically, since an analytical proof is not straightforward under the piecewise-constant assumption.

#### 2.3.3 Midpoint Model

For the time-dependent FOI model, we replace each individual’s exact birth date *b*_*i*_ with the midpoint of their birth interval: 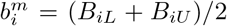. This value is substituted into Eq. 19 to estimate the piecewise-constant FOI over calendar time.

### 2.4 Data

We evaluate our methods on both simulated and real-world serological survey datasets. For simulations, individual-level datasets are generated under three transmission scenarios: constant, age-dependent, and time-dependent FOI. Each dataset includes 2,000 individuals with exact ages drawn uniformly between 0 and 60 years. Binned ages are assigned by grouping individuals into fixed-width intervals as age bins, and midpoint ages are defined as the midpoint of each bin. In all scenarios, serostatus is sampled from a Bernoulli distribution and the seropositivity is calculated using Eq.1, Eq.9 and Eq.18, respectively. We also analyze two published real-world datasets, including mumps data from the United Kingdom (UK) [23] and chikungunya virus (CHIKV) data from Burkina Faso and Gabon [24].

## 3 Results

### 3.1 Constant FOI: The model not accounting for uncertainty in age generally underestimates the FOI

Figure 2 summarizes the performance of the three models under the constant FOI scenario. Figure 2A shows the histogram of the FOI estimates from a single simulated datasets. When only binned ages are available, the commonly used midpoint model increasingly underestimates the FOI as bin size and 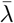 increase. This underestimation occurs because seropositivity increases nonlinearly with age, as illustrated in Figure 1(b), and according to Jensen’s inequality, evaluating it at the midpoint ages gives a higher value than the average seropositivity within that age bin. To match the observed seropositivity, the model compensates by inferring a lower FOI during fitting. A formal proof of this underestimation is provided in §S3.1.

**Figure 2:**
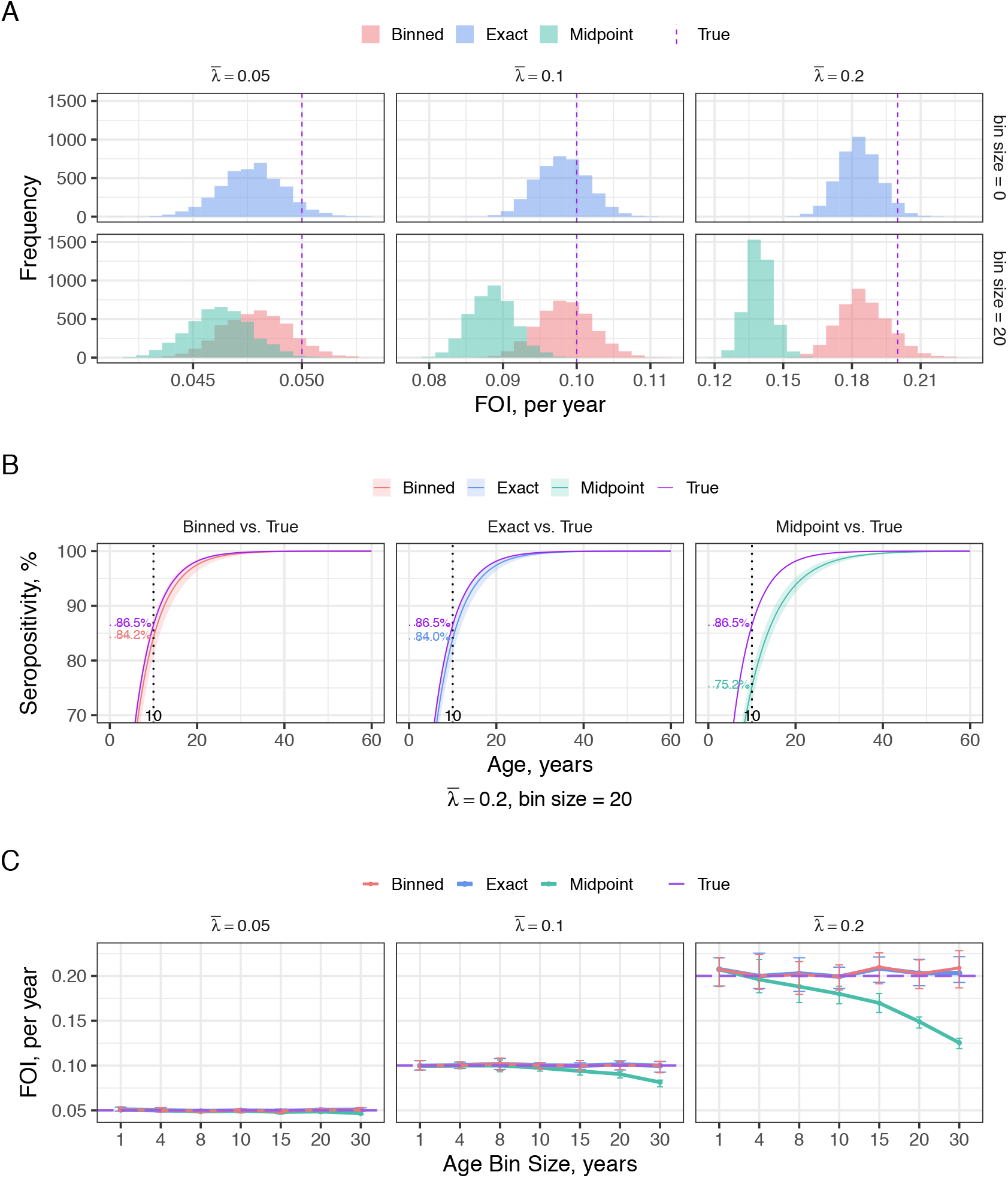
The effect of uncertainty in age on estimates of a constant FOI. The plots compare posterior estimates from the exact (blue), binned (red), and midpoint (green) models against the true values (purple). Panels A and B are based on a single simulated dataset, while Panel C summarizes the mean results from 20 repeated simulations for each combination of 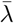 and bin size. **Panel A** shows histograms of the posterior mean estimates of the constant FOI. Each column corresponds to a different true FOI value 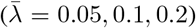, and the rows represent age bin sizes of 0 and 20 years. **Panel B** presents the reconstructed seropositivity curves with 95% credible intervals, obtained using the posterior mean estimates from the bottom-right plot in Panel A (bin size = 20,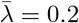). These are compared against the corresponding true seropositivity curves. The dashed vertical line marks age 10, where predicted probabilities are annotated for model comparison. **Panel C** displays posterior mean estimates of the constant FOI with 2.5%-97.5% credible intervals across different age bin sizes, based on 20 repeated simulations for each FOI value. The exact and binned models yield nearly identical estimates, which leads to substantial overlap of the corresponding curves.

Figure 2B compares the reconstructed seropositivity curves using the FOI estimates from the bottom-right plot in Figure 2A. As expected, the exact model performs best, closely followed by the binned model. The midpoint model, however, can deviate by up to 10% from the true seropositivity at certain ages.

Figure 2C presents mean estimates under 20 repeated simulations for each FOI value. The results demonstrate the stability and accuracy of the binned model under repeated sampling and further highlight the divergence between the midpoint and binned models.

### 3.2 Age-dependent FOI: If study participants’ exact ages are unavailable, the model accounting for uncertainty in age produces more reliable FOI estimates

Figure 3 compares the estimated age-dependent FOIs using the three models. Rows correspond to different transmission intensities, with peak intensities specified to be to 0.05, 0.1 and 0.2, and columns denote distinct age bin sizes.

**Figure 3:**
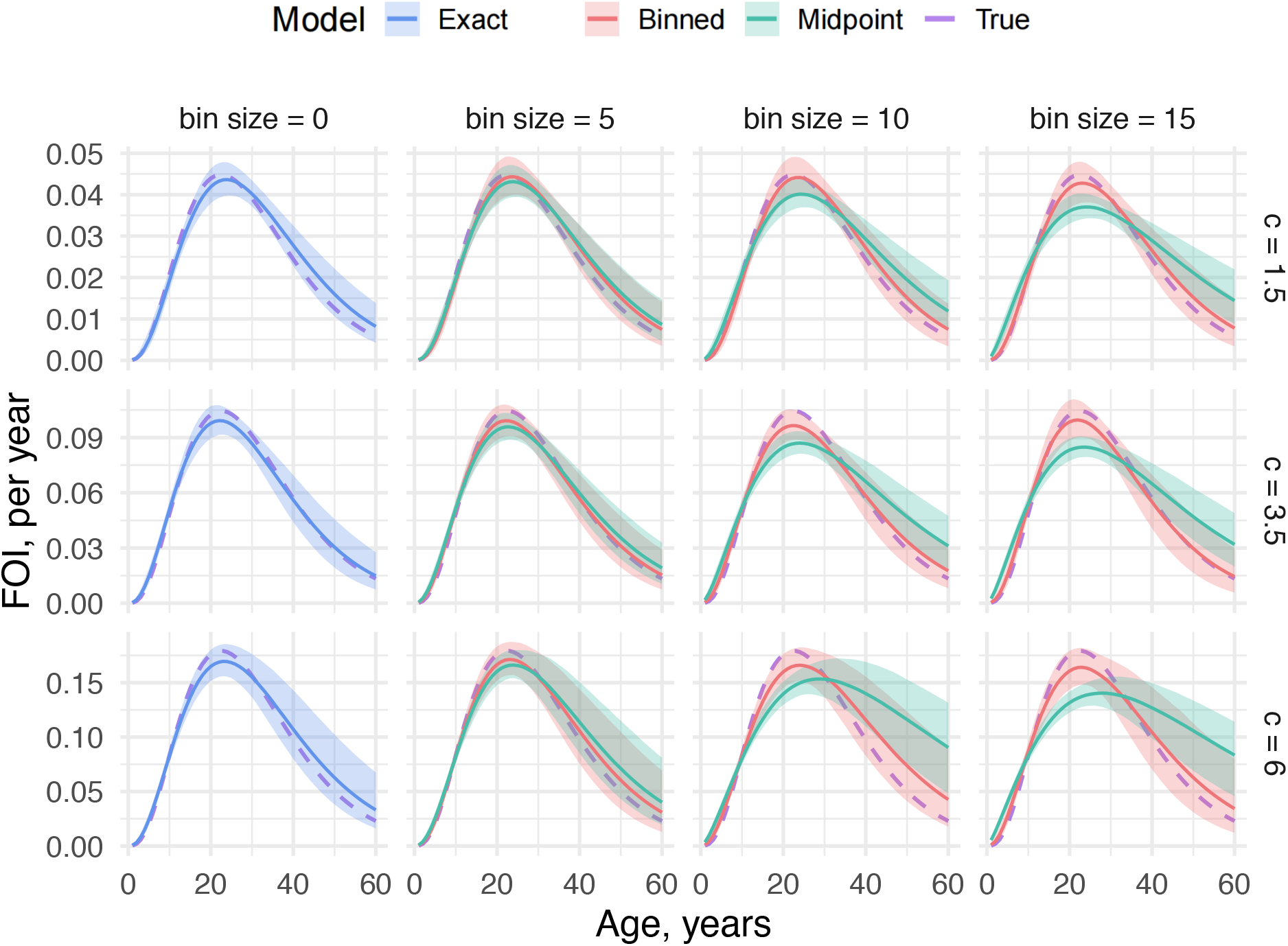
Assessing the reliability of modal age-dependent models in presence of uncertainty in age (*µ* = 30 and *σ* = 15). Each row and each column corresponds to a different value of the scaling constant *c* and bin width used in model fitting. For each combination, the true FOI (purple) is compared against the posterior mean estimates from the exact model (blue), binned model (red) and the midpoint model (green), with the corresponding shaded ribbons indicating 2.5%-97.5% credible intervals.

The exact model generally recovers the true FOI across all transmission levels. As bin size increases, both the midpoint and binned models begin to deviate from the truth, with the midpoint model showing more considerable bias. As discussed in §S3.2, this bias stems from two effects inherent in the midpoint model. The forward-process bias represents the fact that, when only age bins are available, seropositivity evaluated at the midpoint of a bin does not equal the average seropositivity over the bin. During model fitting, this mismatch induces a compensating bias, with the midpoint model adjusting its parameter estimates in a way that pushes the inferred FOI towards a wider and flatter shape.

To assess the generality of these results, we performed a sensitivity analysis in §S3.3 by varying gamma parameters, with *σ >* 5 in all simulations to ensure stable convergence. Results confirm that the binned model estimates remain centered around the true FOI without incurring additional computational cost, which supports its use when only binned age data are available.

#### 3.2.1 Real Data: Mumps Virus

Mumps is typically a childhood infection, with the highest risk in children under 10 years old. Following infection, individuals generally acquire lifelong immunity, consistent with our assumption of no seroreversion. We applied age-dependent models to a dataset collected in 1986–1987 in the UK from individuals aged 1–44 years [23]. Ages are recorded in integer years. The original analysis reported a sharply peaked FOI in young children, supporting the use of a modal age-dependent FOI.

Using the same age groupings as in the original study (1–9, 10–19, and ≥20 years), the resulting FOI and reconstructed seropositivity curves are shown in Figure 4. Since the true FOI is unobservable, the exact model serves as the gold standard. Both the midpoint and binned models produce similar results, though the binned model aligns more closely with the exact model in younger age groups. This is likely because seroprevalence reaches nearly 100% by age 10, meaning FOI is most informative in early childhood, and age binning has limited impact beyond that point.

**Figure 4:**
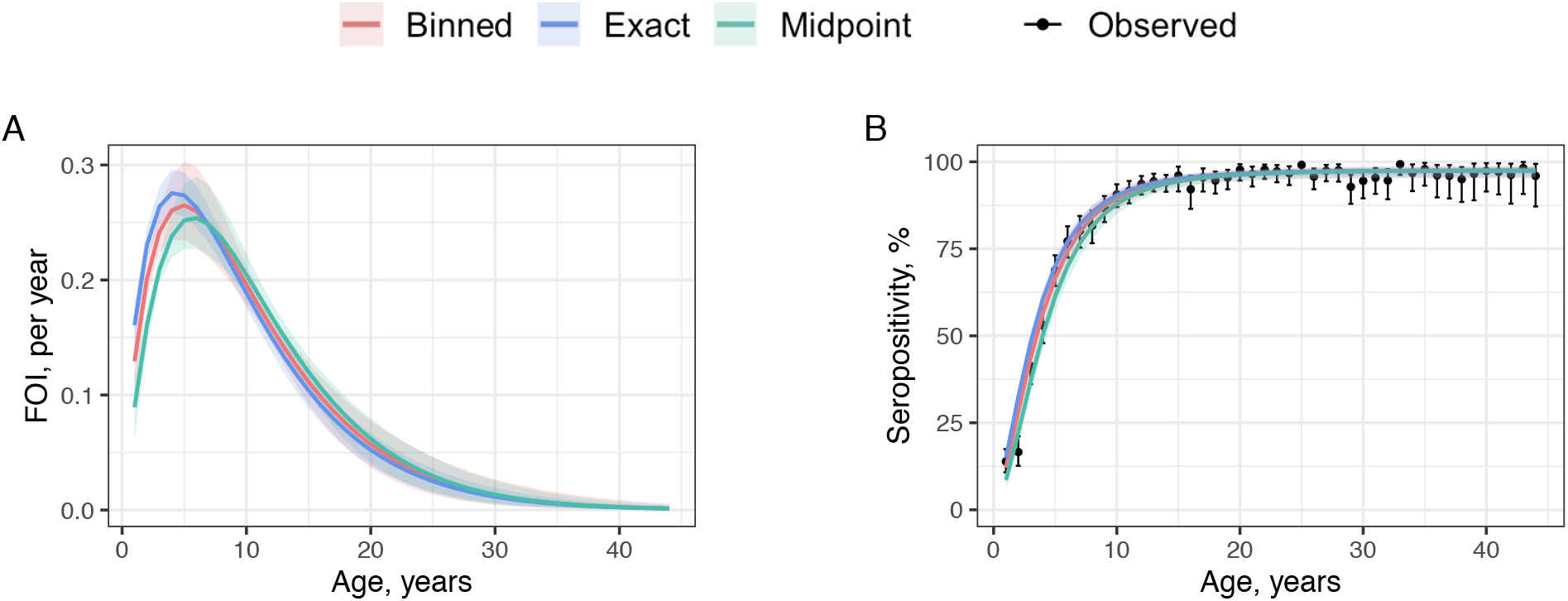
Age-dependent model estimates using mumps virus serological data [23]. **Panel A** shows the posterior mean of the annual FOI estimated using the exact (blue), midpoint (green) and binned (red) models with the 2.5%-97.5% credible intervals. **Panel B** shows the fitted seropositivity curves (lines) reconstructed using the posterior mean of the estimates, compared with observed seropositivity by age (black points showing posterior medians with 2.5%–97.5% Bayesian credible intervals from a Beta(1,1) posterior). Ages are grouped into 1-9, 10-19 and *≥*20 years when using midpoint and binned models.

### 3.3 Time-dependent (piecewise-constant) FOI: It is hard to determine a *priori* how accounting for uncertainty in age will affect estimates

Figure 5 compares time-dependent FOI estimates from exact, midpoint, and binned models under a piecewise-constant assumption that allows flexible representation of real-world transmission patterns, including outbreaks, multiple epidemics, and endemic settings. Simulated data are generated assuming the FOI is constant within 20-year intervals, and the same intervals are used for estimation. This represents an idealized situation. In reality, the true intervals over which the FOI is constant are unknown. Additional transmission patterns and the impact of mismatches between simulation and estimation intervals are explored in §S3.4.

**Figure 5:**
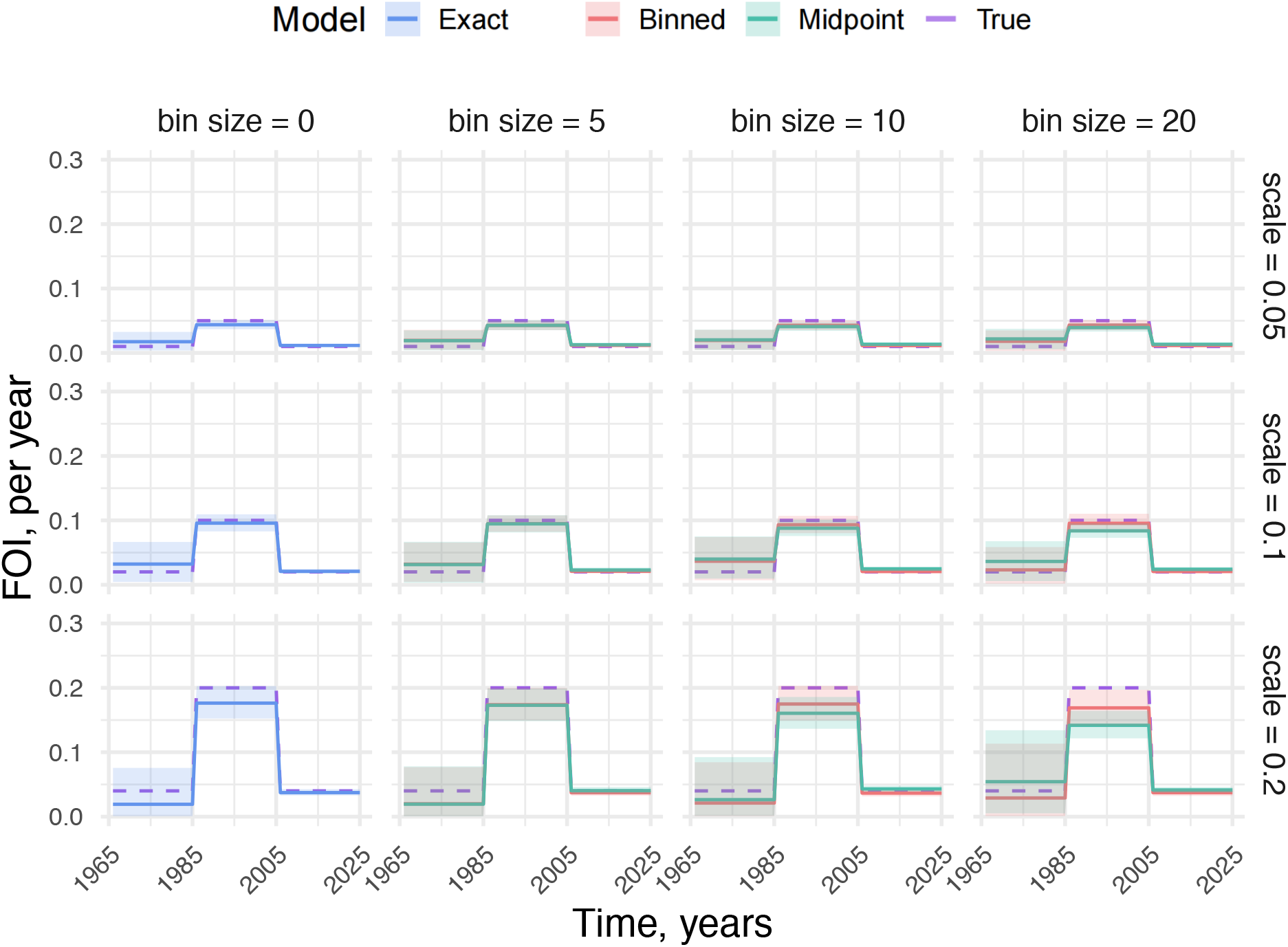
The impact of uncertainty in age on temporally varying FOI estimates. An example set of time-dependent FOI estimates assuming the FOI is piecewise-constant across periods of 20 years. Each row corresponds to a different scale that controls the magnitude of a single FOI spike, and each column to a different age bin size. The true FOI (purple dashed) is compared against estimates from the exact model (blue), binned model (red), and midpoint model (green), with shaded ribbons indicating 2.5%-97.5% credible intervals.

Across all scenarios, the exact model yields the most accurate estimates. However, the effects of age binning on the binned and midpoint model estimates are hard to determine a *priori* and is sensitive to historical transmission risk. This highlights that the bias introduced by age binning is inherently context-dependent and often hard to intuit.

We see in Figure 5 that both the binned and midpoint models tend to perform worse when the underlying FOI is high and the bin size increases, with wider uncertainty particularly for earlier years. This likely reflects that the serostatus of older individuals carries information about transmission, including both earlier and recent years, while younger cohorts only inform about the recent transmission history.

Knowing these variabilities reinforces the value of incorporating uncertainty in age explicitly into the model. Although accounting for this uncertainty does not necessarily guarantee a systematically better fit, it ensures that the inference process remains grounded in the actual structure of the data, rather than imposing unjustified simplifications.

#### 3.3.1 Real Data: Chikungunya virus

To evaluate performance under realistic settings, we apply the midpoint and binned models to a CHIKV dataset, which typically induces long-lasting immunity after infection. The dataset, originally analyzed in [5] and sourced from [24], includes cross-sectional samples from Burkina Faso and Gabon collected in 2015, covering individuals aged between 1-55 years grouped into 5-year age bins.

Using the same prior distributions specified in the previous study [5] (Table S1), we estimate the FOI using the 5-year age bins and the wider 10-year bins. With 5-year bins, both models produce similar FOI estimates, and seropositivities closely match the observed data (Figure 6A-B). As observed in §S3.4, discrepancies between models increase as age bin size grows. Consistent with this, when the analysis is repeated using 10-year bins (Figure 6C-D), differences become evident: the midpoint model tends to underestimate seropositivity, whereas the binned model continues to reproduce the data closely.

**Figure 6:**
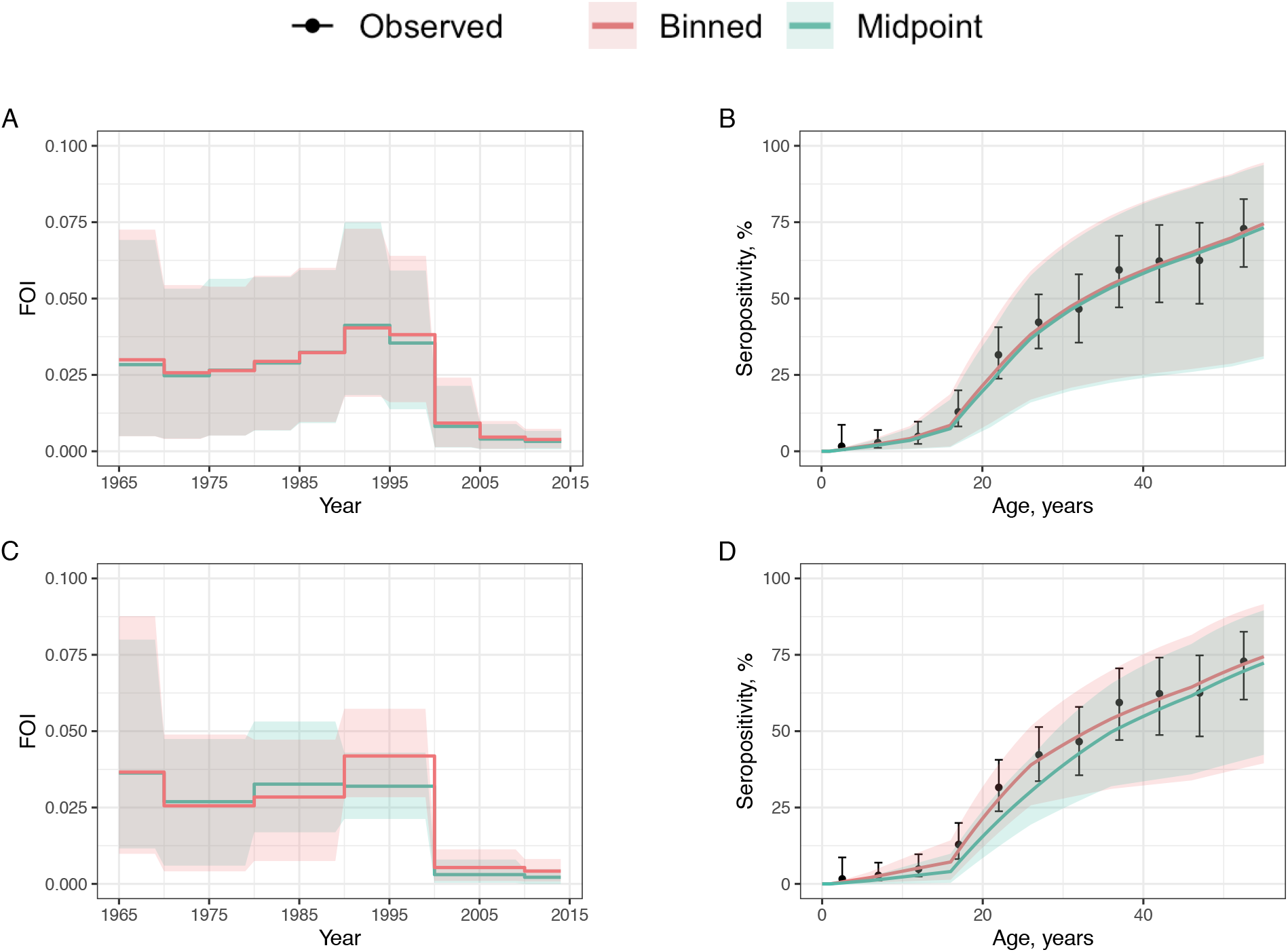
Time-dependent model estimates using CHIKV serological data [24]. **Panels A&C** show the posterior mean of the annual FOI estimated using the midpoint (green) and binned (red) models with the 95% credible intervals. **Panels B&D** show the fitted seropositivity curves (lines) compared with observed seropositivity by age (black points and 95% confidence intervals). The top row presents results using 5-year age bins, while the bottom row shows estimates when ages are grouped into wider 10-year bins.

## 4 Discussion

This study investigates the impact of uncertainty in study participants’ ages in serocatalytic models on FOI estimation. We develop a Bayesian framework that accounts for this uncertainty and compare the results with both inference based on exact ages, treated as a gold standard, and commonly used approaches that assign individuals within an age group to a single midpoint age when exact ages are unavailable. In all the scenarios considered in this study, our results support the use of the exact ages when available, which remains both accurate and computationally efficient. When age information is available only in age groups, approaches that explicitly account for the uncertainty in age generally outperform those that use solely a single midpoint age.

To focus on the impact of uncertainty in participants’ ages on FOI inference, we adopt a set of modeling choices that provide a clear and interpretable baseline and can be straightforwardly extended. We assume that infection-induced mortality is negligible, which is reasonable for viruses like dengue and chikungunya (case fatality ratios (CFR) *<* 0.1%) [25, 26]. For diseases with a high CFR, such as Ebola [27], this assumption could be relaxed by introducing a deceased compartment. We further assume long-lasting immunity after infection, as observed in measles and CHIKV [28, 29], although seroreversion could be incorporated for pathogens with waning immunity or serotype-specific responses. Additional realism could be introduced by accounting for imperfect serological tests [30], allowing FOI to vary jointly with age and calendar time, or modeling the passive transfer and waning of maternal antibodies.

The framework can also accommodate more flexible forms for the FOI profile. While parametric methods like our unimodal gamma model offer interpretability, they cannot capture less regular patterns of variation in FOI. Our piecewise-constant model relies on manually defined cutoffs that are often chosen arbitrarily or to ensure sufficient sample sizes, rather than being learned directly from the data [31]. To address these challenges, future work could explore non-parametric approaches that allow the data to inform the shape and complexity of the FOI profile [32].

Beyond human populations, animal ages are commonly inferred from morphological or physiological proxies, as accurate age determination can be costly, invasive, or infeasible [33, 34, 35, 36]. Even with considerable effort, these estimates are often inaccurate due to individual variability and environmental influences [37, 38, 39, 40]. Given these constraints, wildlife serosurveys tend to report results by age class rather than exact age [34, 41]. In this context, our modeling framework can help reduce practical burdens and improve the reliability of FOI estimation.

Serological data allow insights into infectious disease circulation that are difficult to determine unless routine surveillance is in place. The insights gained from applying serocatalytic models to these data are often used to inform policy, for example, which groups to vaccinate, how to prioritize resources towards areas where there is likely to be a next outbreak. These decisions can hinge on marginal differences in inferred quantities. In our study, we highlight that neglecting the uncertainty in study participants’ age can lead to arbitrary shifts in the inferred FOIs and that these differences are hard to intuit a *priori*. As these outputs can be used to inform policy, we recommend using our model which can more adequately handle this source of uncertainty.

## Acknowledgments

The authors thank James Hay for insightful comments on uncertainty in age in wildlife serosurveys, and the Statistical Epidemiology groups in the Department of Statistics at the University of Oxford and the Pandemic Sciences Institute for helpful comments and suggestions. SB would also like to thank Merton College, University of Oxford where she is the Peter J Braam Early Career Research Fellow in Global Wellbeing.

## Code availability

All simulation and inference pipelines used in this study were built using the *targets* package (version 1.8.0) in R (version 4.3.3) and is publicly available at: github.com/ArianaJunjie/SeroUncertaintyPaper. The repository contains scripts for data simulation, model fitting, and visualization.

## Data availability

All simulated datasets can be reproduced using the code provided in the GitHub repository. Openly available real-world datasets analyzed in this study were originally located at: https://academic.oup.com/jrsssc/article/50/3/251/6990725 for the mumps dataset and https://pubmed.ncbi.nlm.nih.gov/35710849/ for the chikungunya virus dataset. No new data were collected for this study.

## References

[1] L. Cattarino, I. Rodriguez-Barraquer, N. Imai, D. A. T. Cummings, and N. M. Ferguson. “Mapping global variation in dengue transmission intensity”. Science Translational Medicine 12.528 (2020), eaax4144.

[2] N. Hozé et al. “RSero: A user-friendly R package to reconstruct pathogen circulation history from seroprevalence studies”. PLOS Computational Biology 21.2 (2025), e1012777.

[3] A. Menezes, S. L. Razafimahatratra, O. Wariri, A. L. Graham, and C. J. E. Metcalf. “Strengthening serological studies: the need for greater geographical diversity, biobanking, and data-accessibility”. Trends in Microbiology 33.11 (2025), pp. 1155–1162.

[4] H. Salje et al. “Reconstruction of 60 years of chikungunya epidemiology in the Philippines demonstrates episodic and focal transmission”. The Journal of Infectious Diseases 213 (2015).

[5] E. Kamau et al. “The mathematics of serocatalytic models with applications to public health data”. Statistics in Medicine 44. 15–17 (2025), e70188.

[6] Centers for Disease Control and Prevention. Teen Newsletter: Chickenpox. Accessed: 2025-07-29. 2023. URL: https://www.cdc.gov/museum/education/newsletter/2023/nov/index.html.

[7] E. Del Fava et al. “Estimating Age-Specific Immunity and Force of Infection of Varicella Zoster Virus in Norway Using Mixture Models”. PLOS ONE 11.9 (2016), e0163636.

[8] World Health Organization. Influenza (seasonal). Accessed: 2025-07-29. 2025. URL: https://www.who.int/news-room/fact-sheets/detail/influenza-(seasonal).

[9] A. Vicco et al. “A scoping literature review of global dengue age-stratified seroprevalence data: estimating dengue force of infection in endemic countries”. eBioMedicine 104 (2024), p. 105134.

[10] N. P. Macchiaverna, G. F. Enriquez, M. S. Gaspe, and R. E. Gürtler. “Human Trypanosoma cruzi infection in the Argentinean Chaco: risk factors and identification of households with infected children for treatment”. Parasites & Vectors 17.1 (2024), p. 41.

[11] A. Vicco et al. “A simulation-based method to inform serosurvey design for estimating the force of infection using existing blood samples”. PLOS Computational Biology 19.11 (2023), e1011666.

[12] V. Cox et al. “Estimating dengue transmission intensity from serological data: A comparative analysis using mixture and catalytic models”. PLOS Neglected Tropical Diseases 16.7 (2022), e0010592.

[13] J. Mossong et al. “Parvovirus B19 infection in five European countries: Seroepidemiology, force of infection and maternal risk of infection”. Epidemiology and Infection 136 (2008), pp. 1059–68.

[14] K. Nakajo and H. Nishiura. “Age-dependent risk of respiratory syncytial virus infection: A systematic review and hazard modeling from serological data”. The Journal of Infectious Diseases 228.10 (2023), pp. 1400–1409.

[15] S. Alam, N. Hazarika, A. Sarmah, and N. Sarmah. “A retrospective study of measles outbreak investigation in North East India”. International Journal of Current Microbiology and Applied Sciences 4 (2015), pp. 399–405.

[16] European Centre for Disease Prevention and Control. Measles and Rubella Monthly Report. Accessed: 2025-07-09. 2025. URL: https://measles-rubella-monthly.ecdc.europa.eu/.

[17] R. D. Govender, M. Hashim, M. Khan, H. Mustafa, and G. Khan. “Global epidemiology of HIV/AIDS: A resurgence in North America and Europe”. Journal of Epidemiology and Global Health 11.3 (2021).

[18] B. Weaver. “The burden of herpes zoster and postherpetic neuralgia in the United States”. The Journal of the American Osteopathic Association 107 (2007), S2–7.

[19] N. Turner et al. “Quantifying the incidence and burden of herpes zoster in New Zealand general practice: A retrospective cohort study using a natural language processing software inference algorithm”. BMJ Open 8 (2018), e021241.

[20] B. Carpenter et al. “Stan: A Probabilistic Programming Language”. Journal of Statistical Software 76.1 (2017), pp. 1–32.

[21] D. Fisman. “Seasonality of viral infections: mechanisms and unknowns”. Clinical Microbiology and Infection 18.10 (2012), pp. 946–954.

[22] W. Qiu et al. “Effect of public health interventions on COVID-19 cases: an observational study”. Thorax 76.8 (2021), thoraxjnl-2020–215086.

[23] C. P. Farrington, M. N. Kanaan, and N. J. Gay. “Estimation of the basic reproduction number for infectious diseases from age-stratified serological survey data”. Journal of the Royal Statistical Society: Series C (Applied Statistics) 50.3 (2002), pp. 251–292.

[24] J. K. Lim et al. “Seroepidemiological reconstruction of long-term chikungunya virus circulation in Burkina Faso and Gabon”. The Journal of Infectious Diseases 227.2 (2023), pp. 261–267.

[25] N. Haider et al. “Global dengue epidemic worsens with record 14 million cases and 9000 deaths reported in 2024”. International Journal of Infectious Diseases 158 (2025), p. 107940.

[26] C. J. Puntasecca, C. H. King, and A. D. LaBeaud. “Measuring the global burden of chikungunya and zika viruses: A systematic review”. PLOS Neglected Tropical Diseases 15.3 (2021), e0009055.

[27] World Health Organization. Ebola disease. https://www.who.int/news-room/fact-sheets/detail/ebola-disease. Accessed: 2025-07-08. 2025.

[28] H. Auerswald et al. “Broad and long-lasting immune protection against various Chikungunya genotypes demonstrated by participants in a cross-sectional study in a Cambodian rural community”. Emerging Microbes & Infections 7.1 (2018), pp. 1–13.

[29] Q. Wang et al. “Long-term measles antibody profiles following different vaccine schedules in China, a longitudinal study”. Nature Communications 14.1 (2023), p. 1746.

[30] N. Alexander, M. Carabali, and J. K. Lim. “Estimating force of infection from serologic surveys with imperfect tests”. PLOS ONE 16.3 (2021), e0247255.

[31] S. Blaizot et al. “Sample size calculation for estimating key epidemiological parameters using serological data and mathematical modelling”. BMC Medical Research Methodology 19.51 (2019).

[32] R. Creswell et al. “A Bayesian nonparametric method for detecting rapid changes in disease transmission”. Journal of Theoretical Biology 558 (2023), p. 111351.

[33] C. W. Severinghaus. “Tooth development and wear as criteria of age in white-tailed deer”. The Journal of Wildlife Management 13.2 (1949), pp. 195–216.

[34] K. Kim and K. Ito. “Targeted sampling reduces the uncertainty in force of infection estimates from serological surveillance”. Frontiers in Veterinary Science 9 (2022), p. 754255.

[35] A. T. Lu et al. “Universal DNA methylation age across mammalian tissues”. Nature Aging 3.8 (2023), pp. 853–869.

[36] W. M. Parker et al. “Evergrowing incisors of diprotodont marsupials record age and life history”. Archives of Oral Biology 165 (2024), p. 106018.

[37] P. S. Gipson, W. B. Ballard, R. M. Nowak, and L. D. Mech. “Accuracy and precision of estimating age of gray wolves by tooth wear”. The Journal of Wildlife Management 64.3 (2000), pp. 752–758.

[38] H. A. Jacobson and R. J. Reiner. “Estimating age of white-tailed deer: Tooth wear versus cementum annuli”. Proceedings of the Annual Conference Southeastern Association of Fish and Wildlife Agencies 43 (1989), pp. 286–291.

[39] J. D. Wehausen, C. J. O’Brien, and D. R. McCullough. “Reliability of tooth cementum rings to age bighorn sheep: A blind test”. California Fish and Wildlife Journal 110.1 (2024).

[40] C. G. Bell et al. “DNA methylation aging clocks: challenges and recommendations”. Genome Biology 20 (2019), p. 249.

[41] S. Fakour, S. Naserabadi, and E. Ahmadi. “The first positive serological study on Rift Valley fever in ruminants of Iran”. Journal of Vector Borne Diseases 54.4 (2017), pp. 348–352.

