## Supplemental Material for "Accounting for uncertainty in participant age in serocatalytic models"

#### S1 Supplementary Material for Introduction

##### S1.1 Seropositivity Expression under Constant FOI Model Given Exact Ages

- $S^b(t)$: proportion of individuals born at time $b$ who are susceptible (seronegative) to the disease at time $t$,
- $X^b(t)$: proportion of individuals born at time $b$ who are seropositive (presence of antibodies) at time $t$,
- $\bar{\lambda}$: constant FOI, independent of age or time.

$$X^b(t) + S^b(t) = 1, \tag{S1}$$

$$\frac{dS^b(t)}{dt} = -\bar{\lambda} S^b(t), \tag{S2}$$

$$\frac{dX^b(t)}{dt} = \bar{\lambda} S^b(t). \tag{S3}$$

Solve Eq. S2 by integrating both sides:

$$\frac{1}{S^b(t)}\, dS^b(t) = -\bar{\lambda}\, dt \quad \xRightarrow{\text{Separation of variables}} \quad \int_b^t \frac{1}{S^b(t')} dS^b(t') = -\bar{\lambda} \int_b^t dt',$$

where $t'$ is a dummy variable and $b$ is the birth time.

$$[\ln |S^b(t')|]_b^t = -\bar{\lambda}[t']_b^t \quad ,$$

$$\ln |S^b(t)| - \ln |S^b(b)| = -\bar{\lambda} t - (-\bar{\lambda} b).$$

Assume everyone is born to be seronegative (susceptible): $S^b(b) = 1$

$$\ln |S^b(t)| = -\bar{\lambda}(t - b),$$

$$S^b(t) = e^{-\bar{\lambda}(t-b)}.$$

From Eq. S1:

$$X(t \mid \bar{\lambda}) = 1 - S^b(t) = 1 - e^{-\bar{\lambda}(t-b)}.$$

Consider an individual born at time $b$. At time $t$, their exact age is $a := t - b$. The probability of being seropositive at this age $a$ in this constant FOI scenario is given by: $X_{\text{exact}}(a \mid \bar{\lambda}) = 1 - e^{-\bar{\lambda}a}$. This is the derivation of the expression of seropositivity under the constant FOI model used in §2.1.

### S2 Supplementary Material for Methods

#### S2.1 Model Inference

All simulation and inference pipelines used in this study are built using the *targets* package (version 1.8.0) [1] in R (version 4.3.3) [2]. Models are run through the *rstan* interface (version 2.32.7) [3] in R. The prior distributions used in this study can be found in Table S1. Each fit use four independent chains with 2,000 iterations per chain (including 1,000 warmup). Convergence diagnostics are performed using $\hat{R} \leq 1.01$ to ensure reliable inference across all simulation scenarios.

In Table S1, for the constant FOI model, we use a weakly informative prior. An upper bound of 10 per year means the probability of an individual remaining susceptible in one unit time (year) is extremely small, equals $\exp(-10)$.

For the age-dependent scenario, we again use weakly informative prior distributions for all parameters. We specify the prior distribution on $\mu$ to match the age range of the simulated dataset so that the peak of infection occurs within the observed ages. The prior distribution for $\sigma$ allows values up to 50 years, meaning that the standard deviation of the gamma-shaped age-dependent FOI can be as large as 50 years, which is wide relative to the 0–60 age range. This allows both narrow and highly spread-out transmission patterns. Finally, $c$ corresponds to the overall magnitude of the FOI. We cap the prior distribution for $c$ at 20 to ensure that the FOI can reach relatively high levels of infection risk.

For the time-dependent FOI model fitted to simulated data, we use the random walk prior distribution, with the standard deviation $\sigma_\lambda$ equal to 0.1 for all simulations, which allows $\lambda_t$ to vary smoothly over time with a moderate degree of temporal variation.

For the time-dependent FOI model fitted to real data, we use prior distributions consistent with those specified in the previous study to make the model estimates comparable.

Table S1: **Prior Distributions.**

| **Model** | **Prior distributions** | **Corresponding Section** |
|---|---|---|
| Constant FOI, $\bar{\lambda}$. | $\bar{\lambda} \sim \text{Uniform}(0, 10)$ | §3.1 |
| Age-dependent FOI on both simulated and real data, parametrized by $\mu$, $\sigma$ and $c$. | $\mu \sim \text{Uniform}(0, 60)$, $\sigma \sim \text{Uniform}(0, 100)$ and $c \sim \text{Uniform}(0, 10)$ | §3.2 and §3.2.1 |
| Time-dependent FOI on simulated data, $\lambda_t$ | $\lambda_1 \sim \text{Uniform}(0, 1)$ and $\lambda_t \sim \text{Normal}(\lambda_{t-1}, \sigma_\lambda)$, where $\sigma_\lambda = 0.1$. | §3.3 |
| Time-dependent FOI on real data, $\lambda_t$. | $\log \lambda_1 \sim \mathcal{N}(-3, 1)$ and $\log \lambda_t \mid \log \lambda_{t-1} \sim \text{Student-}t(\nu, \log \lambda_{t-1}, \sigma)$, where $\sigma \sim \text{Cauchy}(0, 1)$ and $\nu \sim \text{Cauchy}(0, 1)$ | §3.3.1 |

#### S2.2 Piecewise-constant FOI Assumption

A common approach used to capture heterogeneity in FOI across age or time is to model the FOI as piecewise-constant, where it is constant within predefined intervals, such as one-year age bins, while allowing it to vary between intervals. We describe below how this assumption applies under the age-dependent FOI and time-dependent FOI models, respectively.

##### S2.2.1 Age-dependent FOI

In age-dependent FOI models, the probability of being seropositive is calculated based on how long an individual has lived, assuming that infection risk accumulates with age and immune protection is long-lasting. The cumulative probability of individual $i$ being seropositive by age $a_i$ is given by:

$$X_{\text{exact}}(a_i \mid \lambda_a) = 1 - \exp\left(-\int_0^{a_i} \lambda_a \, da\right),$$

where $\lambda_a$ denotes the FOI at age $a$.

To simplify the model, we assume that FOI is piecewise-constant across predefined age intervals. Since summation boundaries must be integers, we round the age $a_i$ up to the nearest integer:

$$A = \lceil a_i \rceil.$$

We then divide the age range $[0, A)$ into $K$ intervals with boundaries:

$$\tau_0 = 0 < \tau_1 < \cdots < \tau_{K-1} = A.$$

We assign constant FOI values $\lambda_l$ to each interval $[\tau_l, \tau_{l+1})$. In this setup, the function $\lambda_a$ is defined as:

$$\lambda_a = \sum_{l=0}^{K-1} \lambda_l \cdot \mathbb{I}\left[a \in [\tau_l, \tau_{l+1})\right],$$

where $\mathbb{I}[\cdot]$ is the indicator function.

Substituting this into the cumulative formula, the integral becomes:

$$X_{\text{exact}}(a_i \mid \lambda_a) = 1 - \exp\left(-\sum_{l=0}^{K-1} \lambda_l \cdot \Delta_l\right),$$

where $\Delta_l = \max\left(0, \min(a_i, \tau_{l+1}) - \tau_l\right)$ represents the portion of the $l$the interval that falls within $[0, a_i)$.

##### S2.2.2 Time-dependent FOI

In time-dependent FOI models, the probability of being seropositive is calculated based on the calendar years over which an individual has been alive. The underlying assumption is that transmission risk varies over time but affects all individuals equally, regardless of age. The cumulative probability of being seropositive by sampling year $T$ for an individual $i$ born in year $b_i$ is given by:

$$X_{\text{exact}}(b_i \mid \lambda_t) = 1 - \exp\left(-\int_{b_i}^{T} \lambda_t \, dt\right),$$

where $\lambda_t$ denotes the FOI in calendar year $t$.

To simplify the model, we assume that FOI is piecewise-constant across predefined calendar-year intervals. Since the FOI bins are defined by integer-valued years, we round the birth date $b_i$ down to the nearest integer year:

$$B = \lfloor b_i \rfloor.$$

We then divide the interval $[B, T)$ into $K$ intervals with boundaries:

$$\tau_0 = B < \tau_1 < \cdots < \tau_{K-1} = T.$$

We assign constant FOI values $\lambda_l$ to each interval $[\tau_l, \tau_{l+1})$. In this setup, the function $\lambda_t$ is defined as:

$$\lambda_t = \sum_{l=0}^{K-1} \lambda_l \cdot \mathbb{I}\left[t \in [\tau_l, \tau_{l+1})\right],$$

where $\mathbb{I}[\cdot]$ is the indicator function.

Substituting this into the cumulative formula, the integral becomes:

$$X_{\text{exact}}(b_i \mid \lambda_t) = 1 - \exp\left(-\sum_{l=0}^{K-1} \lambda_l \cdot \Delta_l\right),$$

where $\Delta_l = \max\left(0, \min(T, \tau_{l+1}) - \max(b_i, \tau_l)\right)$ represents the portion of the $l$th calendar-year interval that overlaps with the individual's lifetime $[b_i, T)$.

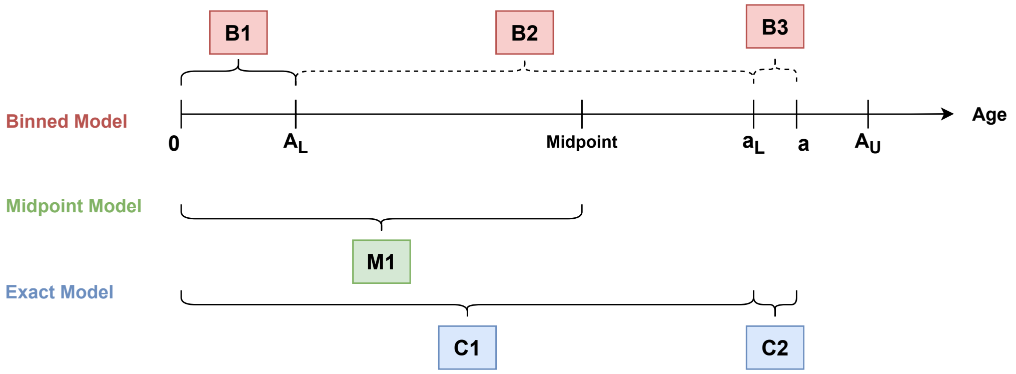


Figure S1: **Illustration of How Age-dependent FOIs are Cumulated.** Different colors correspond to different models, with the binned model shown in red, the midpoint model in green, and the exact model in blue, which are consistent with the color scheme used in the model-specific results. For each model, components of the cumulative FOI calculation are labeled as B1–B3 (binned), M1 (midpoint), and C1–C2 (exact). The dashed curly brackets indicate the age interval where we are uncertain about individuals' exact ages due to the age binning in the participants' ages.

Table S2: Notation Definitions and Examples for Age-dependent Model

| Notations | Explanation | Example |
|---|---|---|
| $a_i$ | Exact age of individual $i$ | An individual of age 6.7 years-old |
| $A_{iL}$ | Integer lower boundary of the age interval where individual $i$ falls | For an age bin $[5, 10)$, $A_{iL} = 5$ |
| $A_{iU}$ | Integer upper boundary of the age interval where individual $i$ falls (Not appear in this equation but will be used in subsequent derivations) | For an age bin $[5, 10)$, since 10 is exclusive and the age boundaries can only be integers, $A_{iU} = 9$ |
| $a_{iL}$ | Integer floor of the individual's exact age $a_i$ | For $a_i = 6.7$, $a_{iL} = 6$ |
| $\lambda_l$ | FOI for individuals in the age interval $[l, l + 1)$ | $\lambda_5$ represents the FOI for individuals aged 5 to 6 |

#### S2.3 Piecewise-constant Age-dependent FOI Model

Figure S1 provides a visual breakdown of how FOI is accumulated by age in the three models to calculate seropositivity for individual $i$ of age $a$. The notation used in the piecewise-constant age-dependent FOI model is summarized in Table S2.

##### S2.3.1 Binned Model

In the binned model, this accumulation is decomposed into three components (B1–B3):

- B1: Individuals exposed to the disease from birth (age 0) up to the lower boundary of age $A_L$. This part is independent of the exact age $a$, as it applies to anyone with the specific $A_L$.
- B2: This is the contribution from $A_L$ to the integer floor of the exact age ($a_L$). Importantly, $a_L$ is dependent on $a$, this breakdown will help in the later derivation of the seropositivity expression in Eq. S6.
- B3: This part represents the remaining fractional contribution from the $a_L$ to $a$. This part also depends on the unobserved exact age.

The cumulative probability of being seropositive at age $a$ can be written as below, where each part corresponds to the region in Figure S1.

$$P_{\text{bin}}(Y_i = 1 \mid a_i, \lambda_a) = 1 - \exp\left( - \Big[ \underbrace{\sum_{l=0}^{A_{iL}-1} \lambda_l}_{\boxed{\text{B1}}} + \underbrace{\sum_{l=A_{iL}}^{a_{iL}-1} \lambda_l}_{\boxed{\text{B2}}} + \underbrace{\lambda_{a_{iL}}(a_i - a_{iL})}_{\boxed{\text{B3}}} \Big] \right). \tag{S4}$$

Dashed lines in the figure corresponding to B2 and B3 indicate components dependent on the exact age, which is unobserved, and hence to account for this age uncertainty, we marginalize over the age interval $[A_L, A_U)$:

$$\begin{aligned} X_{\text{bin}}(A_{iL}, A_{iU} \mid \lambda_a) &= P_{\text{bin}}(Y_i = 1 \mid A_{iL}, A_{iU}, \lambda_a), \\ &= (\frac{1}{A_{iU} - A_{iL}}) \int_{A_{iL}}^{A_{iU}} P_{\text{bin}}(Y_i = 1 \mid a_i, \lambda_a)\, da, \\ &= (\frac{1}{A_{iU} - A_{iL}}) \int_{A_{iL}}^{A_{iU}} 1 - C_i \cdot \int_{A_{iL}}^{A_{iU}} \left[ \exp\left( - \sum_{l=A_{iL}}^{a_L - 1} \lambda_l \right) \cdot \exp\left(-\lambda_{a_L} a\right) \cdot \exp\left(\lambda_{a_L} a_L\right) \right] da. \end{aligned} \tag{S5}$$

where $C_i = \exp\left( - \sum_{l=0}^{A_{iL}-1} \lambda_l \right)$.

Under the piecewise-constant assumption, the binned model reintroduces computational complexity due to the term, $\sum_{l=A_{iL}}^{a_L-1} \lambda_l$, making the integral difficult to solve. Fortunately, we can use a trick to calculate this integration. This is to integrate this probability from $A_{iL}$ to $A_{iU}$ by decomposing the term into sub-intervals $[k-1, k)$. Within each sub-interval, the individual's age $a$ satisfies $a_{iL} = k$, and the summation $\sum_{l=A_{iL}}^{a_{iL}-1} \lambda_l$ becomes constant since the FOI are assumed to be constant within 1 year of age.

$$\begin{aligned} X_{\text{bin}}(A_{iL}, A_{iU} \mid \lambda_a) &= 1 - (\frac{C_i}{A_{iU} - A_{iL}}) \cdot \sum_{k=A_{iL}}^{A_{iU}} \int_{k}^{k+1} \exp\left( - \sum_{l=A_{iL}}^{k-1} \lambda_l \right) \cdot \exp\left(-\lambda_k a\right) \cdot \exp\left(\lambda_k k\right)\, da, \\ &= 1 - (\frac{C_i}{A_{iU} - A_{iL}}) \cdot \sum_{k=A_{iL}}^{A_{iU}} \exp\left( - \sum_{l=A_{iL}}^{k-1} \lambda_l \right) \cdot \exp\left(\lambda_k k\right) \int_{k}^{k+1} \exp\left(-\lambda_k a\right)\, da, \\ &= 1 - (\frac{C_i}{A_{iU} - A_{iL}}) \sum_{k=A_{iL}}^{A_{iU}} \left[ \exp\left( - \sum_{l=A_{iL}}^{k-1} \lambda_l \right) \frac{1}{\lambda_k} \left[1 - \exp(-\lambda_k)\right] \right]. \end{aligned} \tag{S6}$$

Similarly, the likelihood of being seronegative follows as:

$$\begin{aligned} P_{\text{bin}}(Y_i = 0 \mid A_{iL}, A_{iU}, \lambda_a) &= \int_{A_L^i}^{A_U^i} 1 - X_{\text{bin}}(A_{iL}, A_{iU} \mid \lambda_a)\, da, \\ &= C_i \sum_{k=A_{iL}}^{A_{iU}} \left[ \exp\left( - \sum_{l=A_{iL}}^{k-1} \lambda_l \right) \frac{1}{\lambda_k} \left[1 - \exp(-\lambda_k)\right] \right]. \end{aligned} \tag{S7}$$

##### S2.3.2 Exact Model

Given exact ages, Figure S1 breaks the probability of being seropositive into two components as shown in: the cumulative FOI up to the integer floor of the exact age (C1), and the partial contribution due to the fractional part between the exact age and its integer floor (C2). Since the individuals' exact ages $a$ are known in this scenario, $a_{iL}$ is a constant for individual $i$, which removes the additional uncertainty that arises in binned ages.

$$\begin{aligned} X_{\text{exact}}(a_i \mid \lambda_a) &= 1 - \exp\left( - \int_0^a \lambda_a\, da \right), \\ &= 1 - \exp\left( - \Big[ \underbrace{\sum_{l=0}^{a_{iL}-1} \lambda_l}_{\boxed{\text{C1}}} + \underbrace{\lambda_{a_{iL}}(a_i - a_{iL})}_{\boxed{\text{C2}}} \Big] \right) \end{aligned} \tag{S8}$$

##### S2.3.3 Midpoint Model

For the age-dependent FOI model, we replace each individual's exact ages $a_i$ with the midpoint of their age interval: $a_i^m = (A_{iL} + A_{iU})/2$. This value is substituted into Eq. S8 to estimate the piecewise-constant FOI over age. As shown in Figure S1, this midpoint approximation can introduce systematic bias: if an individual's actual age is higher than the midpoint, the model underestimates their seropositivity, and if their age is lower, it overestimates.

##### S2.3.4 Asymptotic Behavior

As the bin width decreases, we examine the asymptotic behavior for selected age cohorts to assess whether seropositivity estimates derived from binned ages converge to those obtained using exact ages. As shown in Figure S2A, the underlying FOI is assumed to follow a gamma-shaped profile, which is used to simulate the serological data. However, the inference procedure presented here assumes a piecewise-constant FOI. Figure S2C demonstrates that the seropositivity curve derived from binned ages consistently converges to that of the exact model as the bin width decreases. An alternative approach that directly estimates the gamma parameters is described in §2.2.

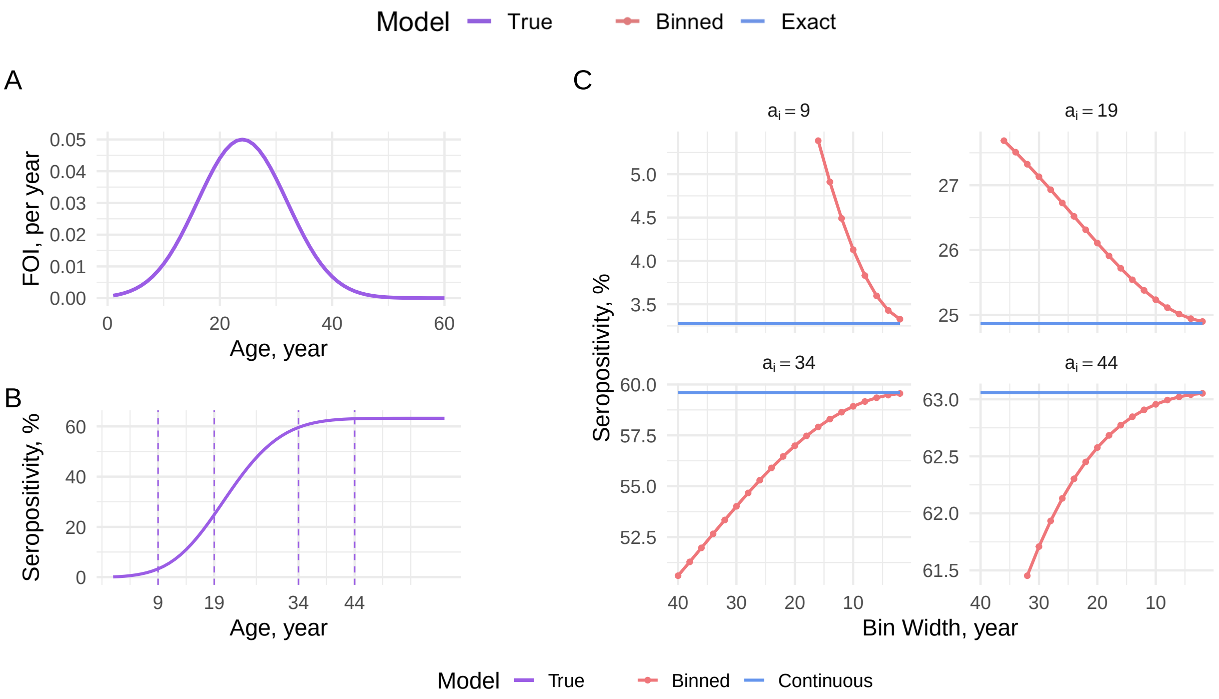


Figure S2: **Asymptotic Behavior under Age-dependent FOI Model.** Purple denotes quantities computed from the true FOI. **Panel A** shows the underlying gamma-shaped age-dependent FOI used to generate the data. **Panel B** shows the resulting seropositivity curve by age, with vertical dashed lines indicating selected ages $(a_i = 9, 19, 34, 44)$ used to examine the asymptotic behavior. **Panel C** illustrates the asymptotic behavior of the estimated posterior mean seropositivity of the binned model (red) compared to the exact model (blue), as the bin width decreases.

#### S2.4 Parametric Age-dependent FOI Model

##### S2.4.1 Asymptotic Behavior

To examine the asymptotic behavior of the binned model as the bin width shrinks, we aim to show that Eq. 13 converges to Eq. 10. Since we use the same prior distributions on $\mu, \sigma$ and c in both models, the key is to prove the following expressions converge at the limit where $A_{iU} \to A_{iL} \to a_i$, i.e.,

$$\lim_{A_{iU} \to A_{iL} \to a_i} \frac{1}{A_{iU} - A_{iL}} \int_{A_{iL}}^{A_{iU}} 1 - \exp\left(-c \cdot \Lambda(a; \mu, \sigma)\right) da = 1 - \exp\left(-c \cdot \Lambda(a_i; \mu, \sigma)\right).$$

Let $h = \frac{A_{iU} - A_{iL}}{2}$, so $A_{iL} = a_i - h$ and $A_{iU} = a_i + h$,

$$\frac{1}{A_{iU} - A_{iL}} \int_{A_{iL}}^{A_{iU}} 1 - \exp\left(-c \cdot \Lambda(a; \mu, \sigma)\right) da = \frac{1}{2h} \int_{a_i - h}^{a_i + h} 1 - \exp\left(-c \cdot \Lambda(a_i; \mu, \sigma)\right) da.$$

Since $1 - \exp\left(-c \cdot \Lambda(a_i; \mu, \sigma)\right)$ is continuous, by the Mean Value Theorem for Integrals, there exists a $\xi_h \in [a_i - h, a_i + h]$ such that:

$$\frac{1}{A_{iU} - A_{iL}} \int_{A_{iL}}^{A_{iU}} 1 - \exp\left(-c \cdot \Lambda(a; \mu, \sigma)\right) da = 1 - \exp\left(-c \cdot \Lambda(\xi_h; \mu, \sigma)\right).$$

As $h \to 0$, $\xi_h \to a_i$, by continuity,

$$\lim_{h \to 0} \frac{1}{A_{iU} - A_{iL}} \int_{A_{iL}}^{A_{iU}} 1 - \exp\left(-c \cdot \Lambda(a; \mu, \sigma)\right) da = 1 - \exp\left(-c \cdot \Lambda(a_i; \mu, \sigma)\right).$$

This proves that the binned model converges to the exact model as the bin width tends to zero.

#### S2.5 Piecewise-constant Time-dependent FOI Model

We assume that the FOI is piecewise-constant within one-year intervals, which defines the smallest units for the integer upper and lower boundaries of individuals' birth time, $B_L$ and $B_U$, used as valid summation limits. If FOI is instead defined over larger intervals, we can easily expand the FOI values by assigning the same values to multiple years. We also assume that the birth time is represented by the unit of date, rather than a precise timestamp, due to data availability and sufficient resolution for our analysis.

What we want to derive from the time-dependent model are the annual probabilities of infection during the time between birth and sampling. *Birth cohorts* refer to groups of individuals born within the same calendar period, typically a year or a range of years. Individuals in the same birth cohort are assumed to share similar infection risk histories, since they were exposed to the same transmission patterns over time. One thing worth noting is that older individuals were born earlier, which creates an inverse relationship between age and birth year. This relationship can be expressed as $B_L = T_L - A_U$ and $B_U = T_L - A_L$, where $T_L$ is the year of the fixed survey date $T$. The boundaries are defined relative to $T_L$ (rather than the exact sampling date $T$) because $B_L$ and $B_U$ are integers denoting integer birth years, which are necessary for use as summation boundaries.

Figure S3 illustrates how the time-dependent FOI, $\lambda_t$, accumulates over calendar time under the three modeling approaches. The notation used for the time-dependent model is summarized in Table 2, and explanations of the breakdown components can be found in Table S3.

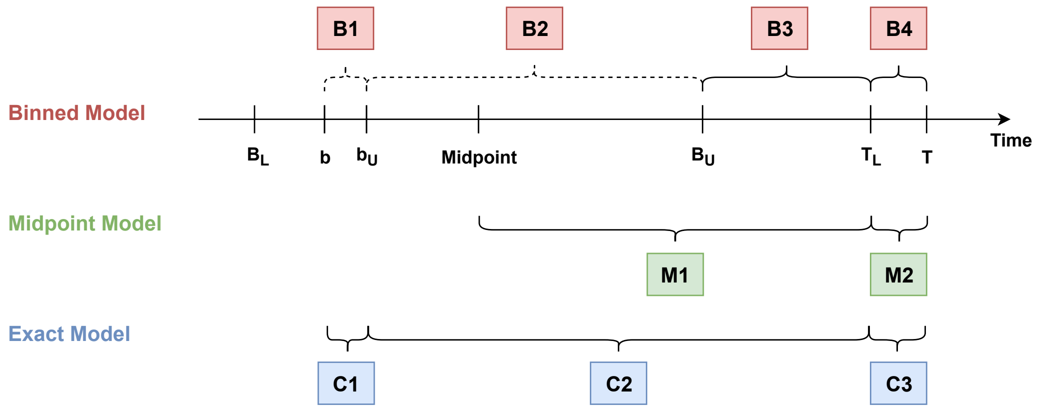


Figure S3: **Illustration of How Time-dependent FOIs are Cumulated.** Different colours correspond to different models, with the binned model shown in red, the midpoint model in green, and the exact model in blue, which are consistent with the colour scheme used in the model-specific results. For each model, components of the cumulative FOI calculation are labeled as B1–B4 (binned), M1–M2 (midpoint), and C1–C3 (exact). The dashed curly brackets indicate the time points where we are uncertain about individuals' birth due to binning in participants' ages.

Table S3: **Decomposition of Accumulated Infection Risk under the Time-dependent FOI Model.** For each modeling approach, the total exposure is expressed as a sum of fractional-year and integer-year components. The decomposition highlights how uncertainty in participants' birth time may affect exposure accumulation under different modeling approaches.

| Region | Interpretation | Notes |
|---|---|---|
| **Binned model** | | |
| B1[a] | Fractional exposure in the birth year, from $b$ to $b_U$. | Summation boundaries are only valid for integer years as defined in Eq. 14, so this part must be added separately. |
| B2[a] | Integer-year exposure between $b_U$ and $B_U$. | This component depends on the unobserved $b$ through $b_U$, separating it is necessary for deriving $P(\lambda_t \mid Y, B_L, B_U)$. |
| B3 | Shared exposure between $B_U$ and $T_L$. | Independent of $b$, common to all individuals in $[B_L, B_U)$. |
| B4 | Fractional exposure in the sampling year, from $T_L$ to $T$. | $T$ may be fractional, so this region is also added separately due to the integer summation boundaries. |
| **Exact model[b]** | | |
| C1 | Same as B1. | Marginalization over $[B_L, B_U)$ is not required since $b$ is observed. |
| C2 | Exposure accumulated over fully observed integer years. | Decomposition by birth interval is not required because $b_U$ is known, given the observed $b$. |
| C3 | Same as B4. | |
| **Midpoint model[b]** | | |
| M1 | Exposure accumulated from midpoint birth date $b^m$ to $T_L$. | Uses $b^m = (B_L + B_U)/2$, so common to all individuals in $[B_L, B_U)$. |
| M2 | Same as B4. | |

[a] Under the binned model, the birth date $b$ is unobserved. The decomposition into B1 and B2 is retained because it is useful for the mathematical derivation of the likelihood.

[b] For the exact and midpoint models, the seropositivity can be equivalently calculated by integrating over the exposure window using Eq. 19, which avoids the integer-year summation boundary. The present decomposition is to visualize differences in accumulated infection risk with the binned model.

##### S2.5.1 Binned Model

Under the binned model, the calculation is divided into four regions, as shown in Figure S3, and the expression of seropositivity is provided in Eq. 14. The definition of $\lambda_t$ is the time-dependent FOI for the interval $[t-1, t)$, so the term, $\lambda_{b_{iU}}$, means the FOI applied to the first partial year after birth, specifically over $[b_i, b_{iU})$. Likewise, $\lambda_{T_L+1}$ denotes the FOI used for the final fractional year of exposure leading up to the survey time $T$, covering the interval $[T_L, T)$.

Since B3 and B4 is independent of the unknown birth date, $b$, we define $C_i = \exp(-\sum_{l=B_{iU}+1}^{T_L} \lambda_l) \cdot \exp(-\lambda_{T_L+1}(T - T_L))$, and then the probability of being seropositive in Eq. 14 can be simplified to:

$$P_{\text{bin}}(Y_i = 1 \mid b_i, \lambda_t) = 1 - \left[ C_i \cdot \exp\left(-\lambda_{b_{iU}}(b_{iU} - b_i)\right) \cdot \exp\left( - \sum_{l=b_{iU}+1}^{B_{iU}} \lambda_l \right) \right]. \tag{S9}$$

To get the marginal posterior distribution of the time-dependent FOI:

$$\begin{aligned} P(\lambda_t \mid \mathbf{Y}, \mathbf{B_L}, \mathbf{B_U}) &\propto P(\lambda_t) \prod_{i=1}^{n} \int_{B_{iL}}^{B_{iU}} P(b|B_{iL}, B_{iU}) \cdot P_{\text{bin}}(Y_i|b, \lambda_t)\, db, \\ &= P(\lambda_t) \prod_{i=1}^{n} (\frac{1}{B_{iU} - B_{iL}}) \int_{B_{iL}}^{B_{iU}} \left[P_{\text{bin}}(Y_i = 1 \mid b_i, \lambda_t)\right]^{Y_i} \left[1 - P_{\text{bin}}(Y_i = 1 \mid b_i, \lambda_t)\right]^{1-Y_i} db. \end{aligned} \tag{S10}$$

Dashed lines in the figure corresponding to B1 and B2 indicate components dependent on the exact birth date which is unobserved, and hence to account for this uncertainty, we marginalize over the birth year interval $[B_L, B_U)$. The probability of being seropositive given the birth interval becomes:

$$\begin{aligned} X_{\text{bin}}(B_{iL}, B_{iU} \mid \lambda_t) &= P_{\text{bin}}(Y_i = 1 \mid B_{iL}, B_{iU}, \lambda_t), \\ &= (\frac{1}{B_{iU} - B_{iL}}) \int_{B_{iL}}^{B_{iU}} P_{\text{bin}}(Y_i = 1 \mid b_i, \lambda_t)\, db, \\ &= (\frac{1}{B_{iU} - B_{iL}}) \int_{B_{iL}}^{B_{iU}} 1 - C_i \cdot \int_{B_{iL}}^{B_{iU}} \left[ \exp\left(-\lambda_{b_U}(b_U - b)\right) \cdot \exp\left( - \sum_{l=b_U+1}^{B_{iU}} \lambda_l \right) \right] db \end{aligned} \tag{S11}$$

As discussed in the previous section, under the piecewise-constant assumption, the binned model reintroduces computational complexity due to the term, $\sum_{l=b_U+1}^{B_{iU}} \lambda_l$ (B2), making the integral difficult to solve. Fortunately, we can use the same trick as in Eq. S6 to calculate this integration. This is to integrate this probability from $B_{iL}$ to $B_{iU}$ by decomposing the term into sub-intervals $[k-1, k)$. In each sub-interval, $b_U = k$, since $\lambda_t$ is assumed to be constant within 1 year, the term $\exp\left(-\sum_{l=k+1}^{B_{iU}} \lambda_l\right)$ is independent of the birth date.

$$\begin{aligned} &\int_{B_{iL}}^{B_{iU}} \exp\left( - \sum_{l=b_U+1}^{B_{iU}} \lambda_l \right) \cdot \exp\left(-\lambda_{b_{iU}}(b_U - b)\right)\, db, \\ &\qquad = \sum_{k=B_{iL}}^{B_{iU}} \int_{k-1}^{k} \exp\left( - \sum_{l=k+1}^{B_{iU}} \lambda_l \right) \cdot \exp(-\lambda_k(k - b))\, db. \end{aligned} \tag{S12}$$

After applying this to Eq. S11, only B1 is left in the integration, which is easy to solve:

$$\begin{aligned} X_{\text{bin}}(B_{iL}, B_{iU} \mid \lambda_t) &= (\frac{1}{B_{iU} - B_{iL}}) \int_{B_{iL}}^{B_{iU}} 1\, db - C_i \sum_{k=B_{iL}}^{B_{iU}} \int_{k-1}^{k} \exp\left( - \sum_{l=k+1}^{B_{iU}} \lambda_l \right) \exp(-\lambda_k(k-b))\, db, \\ &= (\frac{1}{B_{iU} - B_{iL}}) \int_{B_U^i}^{B_L^i} 1\, db - C_i \sum_{k=B_{iL}}^{B_{iU}} \exp\left( - \sum_{l=k+1}^{B_{iU}} \lambda_l \right) \int_{k-1}^{k} \exp(-\lambda_k(k-b))\, db, \\ &= 1 - (\frac{C_i}{B_{iU} - B_{iL}}) \sum_{k=B_{iL}}^{B_{iU}} \left[ \exp\left( - \sum_{l=k+1}^{B_{iU}} \lambda_l \right) \frac{1}{\lambda_k} \left[1 - \exp(-\lambda_k)\right] \right]. \end{aligned} \tag{S13}$$

Similarly, the probability of being seronegative is $1 - X_{\text{bin}}(B_{iL}, B_{iU} \mid \lambda_t)$, so the marginal probability is derived as follows:

$$\begin{aligned} P_{\text{bin}}(Y_i = 0 \mid B_{iL}, B_{iU}, \lambda_t) &= \int_{B_{iL}}^{B_{iU}} 1 - X_{\text{bin}}(b \mid \lambda_t)\, db, \\ &= (\frac{C_i}{B_{iU} - B_{iL}}) \sum_{k=B_{iL}}^{B_{iU}} \left[ \exp\left( - \sum_{l=k+1}^{B_{iU}} \lambda_l \right) \frac{1}{\lambda_k} \left[1 - \exp(-\lambda_k)\right] \right]. \end{aligned} \tag{S14}$$

##### S2.5.2 Asymptotic Behavior

Due to the additional piecewise-constant assumption introduced for the time-dependent FOI model, deriving an analytical solution that directly implies the behavior of the exact model is challenging. Hence, we plot this behavior numerically in Figure S4 by computing the probability across different birth bin widths. We set the exact birth of individuals to be the midpoint of the birth interval, such that the midpoint model yields the same value as the exact model, and then we examine how the binned estimate changes as the bin shrinks symmetrically toward the midpoint. As the underlying FOI is unknown in practice, we tend to examine whether the binned estimate approaches the exact model as the bin width decreases, as exact model gives a gold standard for comparison.

As expected, in Figure S4, the binned seropositivity converges toward the exact model as the bin width approaches 0. Interestingly, the direction of this convergence varies across birth cohorts: for some cohorts. To further explore this phenomenon, we derive the mechanism underpinning the observed convergence patterns mathematically as follows.

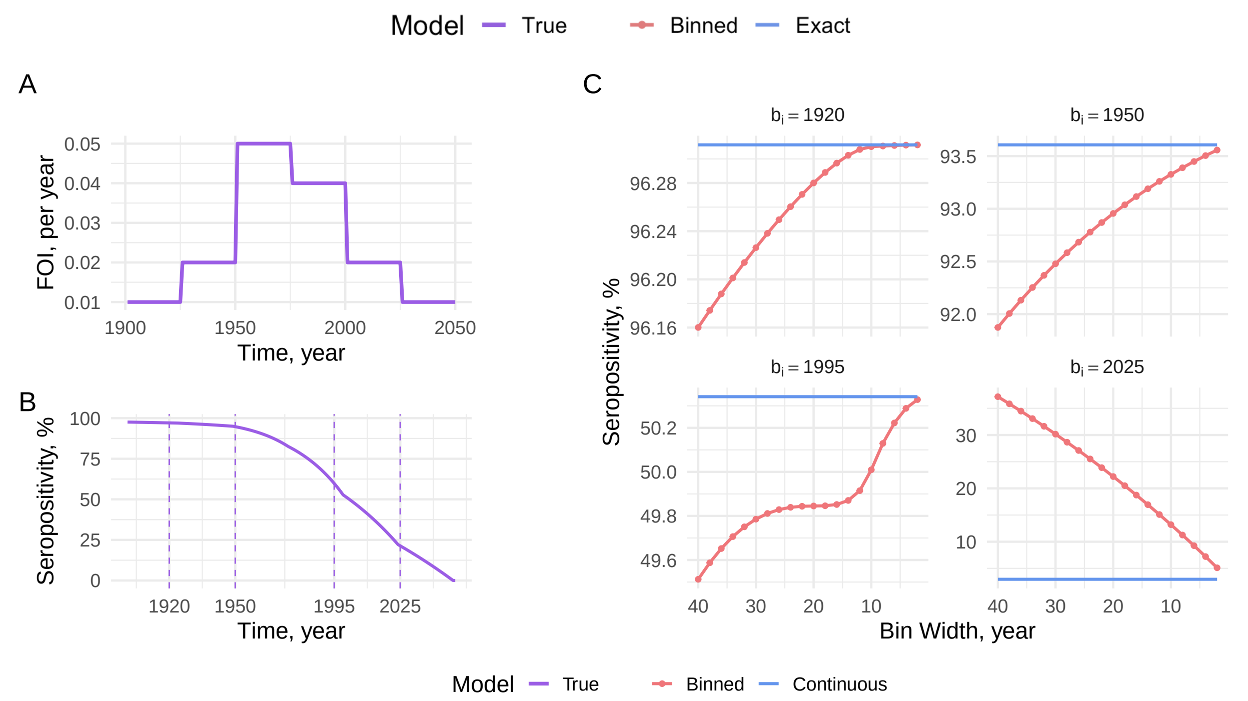


Figure S4: **Asymptotic Behavior of Time-dependent FOI Model.** Purple denotes quantities computed from the true FOI. **Panel A** shows the underlying piecewise-constant time-dependent FOI used to generate the data. **Panel B** shows the resulting seropositivity curve by age, with vertical dashed lines indicating selected ages ($b_i = 1920, 1950, 1995, 2025$) used to examine the asymptotic behavior.**Panel C** illustrates the asymptotic behavior of the binned model (red) compared to the exact model (blue), as the bin width decreases.

Since the prior distribution for $\lambda_t$ is the same between binned and exact models, the key is to show how the FOI accumulates over time. Specifically, the binned model calculates the cumulative FOI by averaging the integral over an age interval:

$$P_{\text{bin}}(B_{iL}, B_{iU} \mid \lambda_t) = \frac{1}{B_{iU} - B_{iL}} \int_{B_{iL}}^{B_{iU}} X_{\text{exact}}(b_i \mid \lambda_t)\, db_i,$$

whereas the exact model evaluates it at an exact birth date (Eq. 18):

$$X_{\text{exact}}(b_i \mid \lambda_t) = 1 - \exp\left(-\int_{b_i}^{T} \lambda_t\, dt\right).$$

The birth interval symmetrically centers around the midpoint $b_i^m$, i.e.,

$$B_{iL} = b_i^m - h, \quad B_{iU} = b_i^m + h,$$

so the bin width is $2h$.

$$P_{\text{bin}}(b_i^m, h \mid \lambda_t) = \frac{1}{2h} \int_{b_i^m - h}^{b_i^m + h} X_{\text{exact}}(b \mid \lambda_t)\, db$$

We now differentiate $X_{\text{bin}}(b_i^m, h \mid \lambda_t)$ with respect to $h$ to derive how it changes with bin width:

$$\frac{dP_{\text{bin}}(b_i^m, h \mid \lambda_t)}{dh} = \frac{d}{dh}\left(\frac{1}{2h} \int_{b_i^m - h}^{b_i^m + h} X_{\text{exact}}(b \mid \lambda_t)\, db\right),$$

where we define $I(h) = \int_{b_i^m - h}^{b_i^m + h} X_{\text{exact}}(b \mid \lambda_t)\, db$

Leibniz's Rule is defined as follows:

$$\frac{d}{dh} \int_{\alpha(h)}^{\beta(h)} f(b, h)\, db = f(\beta(h), h)\frac{d\beta}{dh} - f(\alpha(h), h)\frac{d\alpha}{dh} + \int_{\alpha(h)}^{\beta(h)} \frac{\partial f(b, h)}{\partial h}\, db.$$

We apply Leibniz's Rule to the integral $I(h)$, where $\alpha(h) = b_i^m - h; \beta(h) = b_i^m + h; X_{\text{exact}}(b \mid \lambda_t) = f(b,h)$. Since $X_{\text{exact}}(b \mid \lambda_t)$ does not dependent on $h$, $\frac{\partial f(b,h)}{\partial h} = 0$. So the derivative simplifies to:

$$\frac{d}{dh}\int_{b_i^m-h}^{b_i^m+h} X_{\text{exact}}(b \mid \lambda_t)\, db = X_{\text{exact}}(b_i^m + h \mid \lambda_t)\frac{d(b_i^m+h)}{dh} - X_{\text{exact}}(b_i^m - h \mid \lambda_t)\frac{d(b_i^m-h)}{dh}.$$

The derivatives of the limits are $\frac{d(b_i^m+h)}{dh} = 1$ and $\frac{d(b_i^m-h)}{dh} = -1$, so the result becomes:

$$\frac{d}{dh}\int_{b_i^m-h}^{b_i^m+h} X_{\text{exact}}(b \mid \lambda_t)\, db = X_{\text{exact}}(b_i^m + h \mid \lambda_t) + X_{\text{exact}}(b_i^m - h \mid \lambda_t).$$

Therefore:

$$\begin{aligned}\frac{dP_{\text{bin}}(b_i^m, h \mid \lambda_t)}{dh} &= \frac{-1}{2h^2}\int_{b_i^m-h}^{b_i^m+h} X_{\text{exact}}(b \mid \lambda_t)\, db + \frac{1}{2h}\left[X_{\text{exact}}(b_i^m + h \mid \lambda_t) + X_{\text{exact}}(b_i^m - h \mid \lambda_t)\right] \\ &= \frac{1}{h}\left[\frac{X_{\text{exact}}(b_i^m + h \mid \lambda_t) + X_{\text{exact}}(b_i^m - h \mid \lambda_t)}{2} - \frac{1}{2h}\int_{b_i^m-h}^{b_i^m+h} X_{\text{exact}}(b \mid \lambda_t)\, db\right]\end{aligned}$$

Now define the average of the endpoints:

$$M(a_i^m, h) := \frac{X_{\text{exact}}(a_i^m + h \mid \lambda_t) + X_{\text{exact}}(a_i^m - h \mid \lambda_t)}{2}.$$

Then:

$$\frac{dP_{\text{bin}}(b_i^m, h \mid \lambda_t)}{dh} = \frac{1}{h}\cdot\left[M(b_i^m, h) - X_{\text{bin}}(b_i^m, h \mid \lambda_t)\right]. \tag{S15}$$

Therefore, the sign of the derivative depends on the difference between the average of the two endpoints $M(b_i^m, h)$ and the bin-averaged estimate $P_{\text{bin}}(b_i^m, h \mid \lambda_t)$.

The Hermite–Hadamard Inequality states that: for a convex function f on $[a, b]$, the following holds:

$$f(\frac{a+b}{2}) \leq \frac{1}{b-a}\int_a^b f(x)\, dx \leq \frac{f(a)+f(b)}{2},$$

so when $X_{\text{exact}}(b_i^m \mid \lambda)$ is convex, the inequality suggests:

$$X_{\text{exact}}(b_i^m \mid \lambda) \leq P_{\text{bin}}(b_i^m, h \mid \lambda_t) \leq M(b_i^m, h).$$

This inequality can be visualized in Figure S5. In Scenario 1, $X_{\text{exact}}(b \mid \lambda_t)$ is concave, which is a case where the Hermite–Hadamard Inequality reverses:

$$X_{\text{exact}}(b_i^m \mid \lambda) \geq P_{\text{bin}}(b_i^m, h \mid \lambda_t) \geq M(b_i^m, h).$$

The binned model underestimate the seropositivity compared to the exact model evaluated at the midpoint. Since the inequality gives $P_{\text{bin}}(b_i^m, h \mid \lambda_t) \geq M(b_i^m, h)$, substitute back into Eq. S15, we have $\frac{dP_{\text{bin}}(b_i^m, h|\lambda_t)}{dh} < 0$, which implies that the binned estimate increases as the bin size shrinks. This explains the behavior in Figure S4C when $b_i = 1920$, 1950 or 1995, which shows that the binned estimate lies below the exact line and gradually converges toward it as the bin width decreases.

In contrast, Scenario 2 corresponds to the case where $X_{\text{exact}}(b_i^m \mid \lambda_t)$ is convex. In this case, the binned model overestimates the seropositivity relative to the exact model at the midpoint. The derivative in Eq. S15 becomes positive in this case. As shown in Figure S4C when $b_i = 2025$, the binned estimate decreases as the bin size shrinks.

In other words, whether the binned model overestimates or underestimates seropositivity at a given midpoint depends on the local curvature of the seropositivity curve generated by the underlying FOI used to simulate the serosurvey.

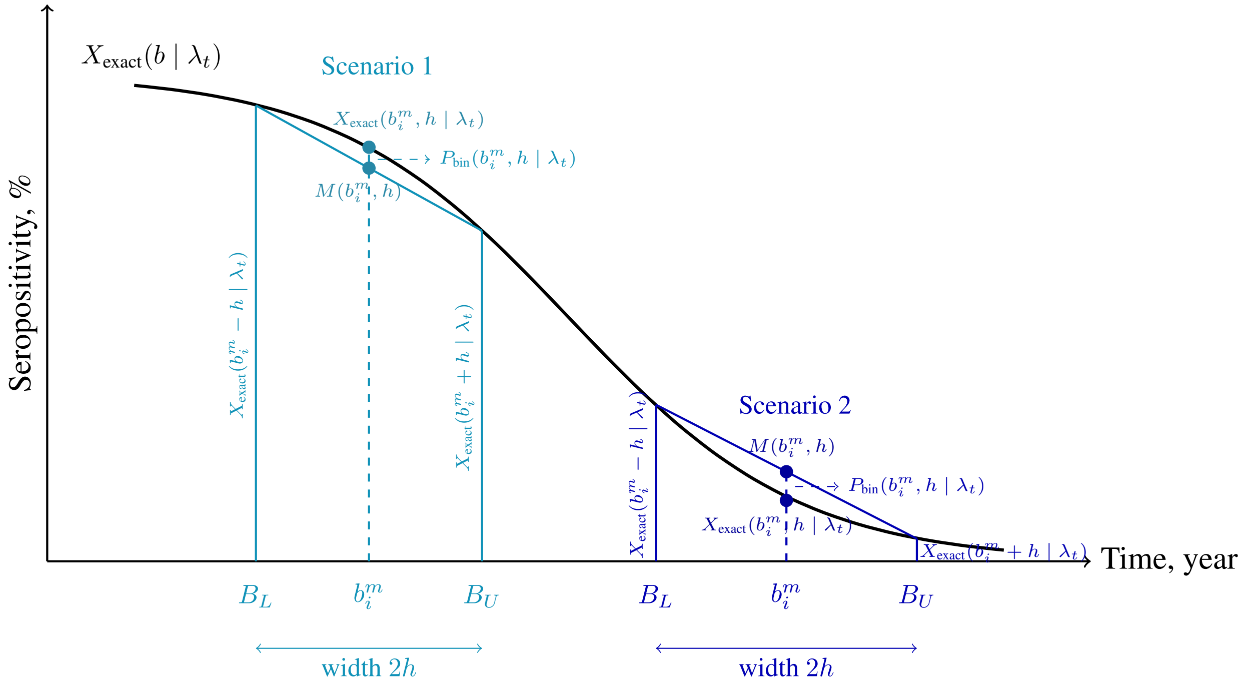


Figure S5: **Illustration of How the Asymptotic Behavior depends on the Curvature of Seropositivity Curves.** This figure illustrates how the bin-averaged seropositivity $P_{\text{bin}}(b_i^m, h \mid \lambda_t)$ relates to the exact seropositivity curve $X_{\text{exact}}(b_i^m \mid \lambda_t)$ at the midpoint birth as the bin width $(2h)$ changes. Two scenarios are shown: in Scenario 1 (light blue), the curve is concave, and in Scenario 2 (dark blue), the curve is convex.

### S3 Supplementary Material for Results

#### S3.1 Proof that the Midpoint Model Always Underestimates the Constant FOI

Let $\bar{\lambda} > 0$ be the constant FOI. Consider an age bin from $A_{iL}$ to $A_{iU}$ of width $2h = A_{iU} - A_{iL}$ and midpoint $a_i^m = \frac{A_{iL}+A_{iU}}{2}$.

The midpoint model assumes all individuals in the bin have age $m_i$, so the probability of being seropositive:

$$X_{\text{mid}}(a_i^m \mid \bar{\lambda}) = 1 - e^{-\bar{\lambda} a_i^m}. \tag{S16}$$

The binned model integrates the seropositivity probability over the interval $[A_{iL}, A_{iU}]$:

$$X_{\text{bin}}(A_{iL}, A_{iU} \mid \bar{\lambda}) = \frac{1}{2h} \int_{A_{iL}}^{A_{iU}} \left(1 - e^{-\bar{\lambda} a}\right) da = 1 - \frac{1}{2h} \int_{A_{iL}}^{A_{iU}} e^{-\bar{\lambda} a}\, da. \tag{S17}$$

Evaluate the integral:

$$\int_{A_{iL}}^{A_{iU}} e^{-\bar{\lambda} a}\, da = \left[-\frac{1}{\bar{\lambda}} e^{-\bar{\lambda} a}\right]_{A_{iL}}^{A_{iU}} = \frac{1}{\bar{\lambda}} \left(e^{-\bar{\lambda} A_{iL}} - e^{-\bar{\lambda} A_{iU}}\right).$$

Thus,

$$X_{\text{bin}}(A_{iL}, A_{iU} \mid \bar{\lambda}) = 1 - \frac{1}{2h} \cdot \frac{1}{\bar{\lambda}} \left(e^{-\bar{\lambda} A_{iL}} - e^{-\bar{\lambda} A_{iU}}\right) = 1 - \frac{e^{-\bar{\lambda} A_{iL}} - e^{-\lambda A_{iU}}}{2\bar{\lambda} h}.$$

Compare the two:

$$\Delta = X_{\text{mid}}(a_i^m \mid \bar{\lambda}) - X_{\text{bin}}(A_{iL}, A_{iU} \mid \bar{\lambda}),$$

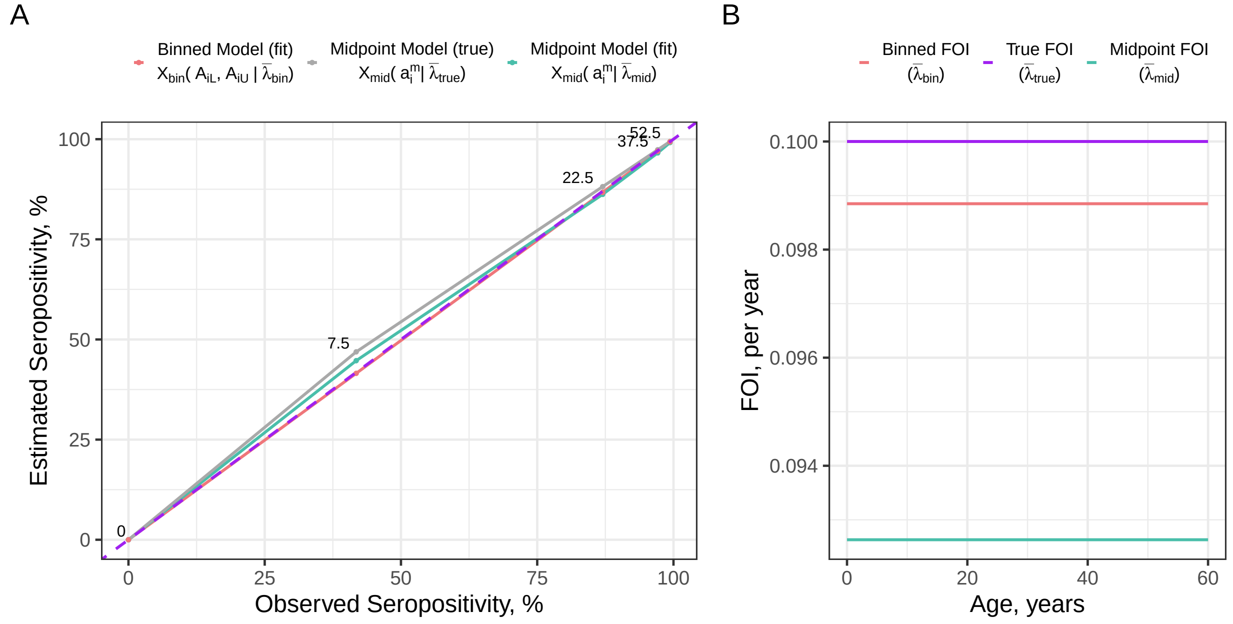


Figure S6: **Illustration of Biases in the Constant FOI Model.** The underlying true FOI used to generate data is $\bar{\lambda}_{\text{true}} = 0.10$, and ages are grouped into 15-year age bins. **Panel A:** Observed versus estimated seropositivity at the bin level (one value per age bin, joined by lines for visual clarity). The legend indicates the seropositivity function representations used to calculate each seropositivity curve, as defined in Eq. S16 and Eq. S17. The numbers shown next to the curves indicate the midpoint age (in years) that corresponds to each age bin. The observed seropositivities on the x-axis represent the proportion seropositive among individuals within each age bin in the data. The purple dashed line indicates perfect agreement, where the estimated seropositivity equals the observed value. The grey line shows the seropositivity evaluated at the midpoint age of each bin under the true constant FOI. The green and red lines show the posterior predictive seropositivity obtained from the midpoint and binned model estimates, respectively. **Panel B:** Constant FOI values that are supplied to Eq. S16 and Eq. S17 to calculate the corresponding seropositivities shown in Panel A: the true FOI (purple), the midpoint model FOI estimate (green), and the binned model FOI estimate (red).

$$
\begin{aligned}
&= \left(1 - e^{-\bar{\lambda} a_i^m}\right) - \left(1 - \frac{e^{-\bar{\lambda} A_{iL}} - e^{-\bar{\lambda} A_{iU}}}{2\bar{\lambda} h}\right), \\
&= \frac{e^{-\bar{\lambda} A_{iL}} - e^{-\bar{\lambda} A_{iU}}}{2\bar{\lambda} h} - e^{-\bar{\lambda} a_i^m}.
\end{aligned}
$$

Now substitute $A_{iL} = a_i^m - h$ and $A_{iU} = a_i^m + h$:

$$
e^{-\bar{\lambda} A_{iL}} = e^{-\bar{\lambda} a_i^m} \cdot e^{\bar{\lambda} h}, \quad e^{-\bar{\lambda} A_{iU}} = e^{-\bar{\lambda} a_i^m} \cdot e^{-\bar{\lambda} h}.
$$

Therefore:

$$
\Delta = \frac{e^{-\bar{\lambda} a_i^m}\left(e^{\bar{\lambda} h} - e^{-\bar{\lambda} h}\right)}{2\bar{\lambda} h} - e^{-\bar{\lambda} a_i^m},
$$

$$
= e^{-\bar{\lambda} a_i^m}\left(\frac{e^{\bar{\lambda} h} - e^{-\bar{\lambda} h}}{2\bar{\lambda} h} - 1\right).
$$

Use the identity $\sinh(x) = \frac{e^x - e^{-x}}{2}$, $e^{\lambda h} - e^{-\lambda h} = 2\sinh(\lambda h)$:

$$
\Delta = e^{-\bar{\lambda} a_i^m}\left(\frac{\sinh(\bar{\lambda} h)}{\bar{\lambda} h} - 1\right).
$$

Since $e^{-\bar{\lambda}a_i^m} > 0$ for all $\bar{\lambda} > 0$, the sign of $\Delta$ depends on:

$$\frac{\sinh(\bar{\lambda}h)}{\bar{\lambda}h} - 1.$$

Since $\sinh(x) > x$ for all $x > 0$, so:

$$\frac{\sinh(x)}{x} > 1 \Rightarrow \Delta > 0.$$

**Conclusion:** For any $\bar{\lambda} > 0$ and $h > 0$,

$$X_{\text{mid}}(a_i^m \mid \bar{\lambda}) > X_{\text{bin}}(a_i^m, h \mid \bar{\lambda}) \Rightarrow X_{\text{mid}}(a_i^m \mid \bar{\lambda}) > X_{\text{bin}}(A_{iL}, A_{iU} \mid \bar{\lambda}). \tag{S18}$$

That is, from Eq. S18, the midpoint model always over-estimates the observed seropositivity. As shown in Figure S6A, the midpoint seropositivity computed using the true FOI (grey) always lies above the diagonal reference line (purple dashed). The binned model, in contrast, explicitly accounts for the age binning uncertainties, and therefore its predicted seropositivity (red) lies close to the diagonal across all bins.

To compensate for this upward bias, inference under the midpoint model tends to underestimate the FOI during model fitting. Figure S6B shows that the FOI estimate from the midpoint model is much smaller than both the true constant FOI and the binned model estimate. As a result, the corresponding posterior predictive seropositivity from the midpoint model (green) shifts downward and becomes closer to the diagonal purple line in Figure S6A.

#### S3.2 Parametric Age-dependent FOI Bias Introduced by Midpoint Model

Here, we quantify the bias introduced by the midpoint model and explore the mechanism underpinning this bias when estimating parametric age-dependent FOIs. We begin by analysing their behaviour in a setting where the underlying FOI curve is treated as known and fixed (i.e., fixed $\mu, \sigma, c$). This follows the same idea as the forward mapping from FOI to seropositivity shown in Figure 1(c)–(d), where exact ages are used to generate the seropositivity curve. Here, we extend that forward process to examine what happens when ages are available only in bins, which allows us to compare the difference between the midpoint and binned seropositivity. We refer to this difference as the forward-process bias.

However, during model fitting, serocatalytic models work in the opposite direction to the forward process (as described in §1). For the midpoint model, since only the number of seropositive individuals and the total number of individuals in each age bin are observed, the model attempts to match the resulting seropositivity pattern by adjusting the FOI parameters. This compensatory adjustment introduces an inverse-process (inference) bias.

Together, the forward-process bias explains why the midpoint likelihood misrepresents the data, and the inverse-process bias explains how the model responds when trying to fit them.

##### S3.2.1 Forward-process Bias

We first compare the midpoint and binned seropositivity expressions assuming the known FOI parameters. Let $\Lambda(a) = \Lambda(a; \mu, \sigma)$, the expressions of the seropositivity curve given the age bin $[A_L, A_U)$ that an individual falls into can be written as $X_{\text{bin}}(A_{iL}, A_{iU} \mid \mu, \sigma, c)$ for the binned model and $X_{\text{mid}}(A_{iL}, A_{iU} \mid \mu, \sigma, c)$ for the midpoint model:

$$X_{\text{mid}}(A_{iL}, A_{iU} \mid \mu, \sigma, c) = 1 - \exp\left(-c \cdot \Lambda(\frac{A_{iL} + A_{iU}}{2})\right). \tag{S19}$$

$$X_{\text{bin}}(A_{iL}, A_{iU} \mid \mu, \sigma, c) = \frac{1}{A_{iU} - A_{iL}} \int_{A_{iL}}^{A_{iU}} [1 - \exp(-c \cdot \Lambda(a))]\, da. \tag{S20}$$

We define $a_i^m = \frac{A_{iL}+A_{iU}}{2}$ and $h = \frac{A_{iU}-A_{iL}}{2}$, so Eq. S19 and Eq. S20 can be simplified as follows:

$$X_{\text{mid}}(a_i^m \mid \mu, \sigma, c) = 1 - \exp(-c \cdot \Lambda(a_i^m)), \tag{S21}$$

$$X_{\text{bin}}(a_i^m, h \mid \mu, \sigma, c) = \frac{1}{2h} \int_{a_i^m - h}^{a_i^m + h} [1 - \exp(-c \cdot \Lambda(a))]\, da. \tag{S22}$$

Expanding $\Lambda(a)$ around $a_i^m$ using second-order Taylor expansion:

$$\Lambda(a) \approx \Lambda(a_i^m) + \Lambda'(a_i^m)(a - a_i^m) + \frac{1}{2}\Lambda''(a_i^m)(a - a_i^m)^2.$$

Hence:

$$\exp\left(-c \cdot \Lambda(a)\right) \approx \exp\left(-c \cdot \Lambda(a_i^m)\right) \cdot \exp\left(-c \cdot \Lambda'(a_i^m)(a - a_i^m) - \frac{1}{2}c \cdot \Lambda''(a_i^m)(a - a_i^m)^2\right).$$

Let $x = a_i - a_i^m$, $\Rightarrow a_i = a_i^m + x_i$ and since $x_i \sim$ Uniform$(-h, h)$, we have:

$$\begin{aligned}
X_{\text{bin}}(a_i^m, h \mid \mu, \sigma, c) &= \frac{1}{2h}\int_{-h}^{h}\left(1 - \exp\left(-c \cdot \Lambda(a_i^m)\right)\exp\left(-c \cdot \Lambda'(a_i^m)x - \frac{1}{2}c \cdot \Lambda''(a_i^m)x^2\right)\right)dx,\\
&= \frac{1}{2h}\int_{-h}^{h}\left[1 - \exp\left(-c\Lambda(a_i^m)\right)f(x)\right]dx \qquad \text{where } f(x) = \exp\left(-c \cdot \Lambda'(a_i^m)x - \frac{1}{2}c \cdot \Lambda''(a_i^m)x^2\right),\\
&= \frac{1}{2h}\int_{-h}^{h} 1\,dx - \frac{1}{2h}\exp\left(-c\Lambda(a_i^m)\right)\int_{-h}^{h} f(x)\,dx,\\
&= 1 - \exp\left(-c\Lambda(a_i^m)\right) \cdot \frac{1}{2h}\int_{-h}^{h} f(x)\,dx.
\end{aligned}$$

Let $B = c \cdot \Lambda'(a_i^m) \cdot x$ and C = $\frac{1}{2}c \cdot \Lambda''(a_i^m) \cdot x^2 \Rightarrow f(x) = \exp(-B - C)$. Using second-order Taylor expansion,

$$\begin{aligned}
f(x) &= \exp(-B - C),\\
&= 1 - (B + C) + \frac{1}{2}(B + C)^2,\\
&= 1 - c \cdot \Lambda'(a_i^m) \cdot x - \frac{1}{2}c \cdot \Lambda''(a_i^m) \cdot x^2 + \frac{1}{2}\left(c \cdot \Lambda'(a_i^m) \cdot x + \frac{1}{2}c \cdot \Lambda''(a_i^m) \cdot x^2\right)^2,\\
&\approx 1 - c \cdot \Lambda'(a_i^m) \cdot x - \frac{1}{2}c \cdot \Lambda''(a_i^m) \cdot x^2 + \frac{1}{2}\left(c \cdot \Lambda'(a_i^m) \cdot x\right)^2.
\end{aligned} \tag{S23}$$

Eq. S23 drops the cross term and the higher order since we are using the second-order approximation.

Since

$$\begin{aligned}
\int_{-h}^{h} 1\,dx &= 2h,\\
\int_{-h}^{h} x\,dx &= 0,\\
\int_{-h}^{h} x^2\,dx &= \frac{2}{3}h^3,
\end{aligned}$$

we have:

$$\begin{aligned}
\frac{1}{2h}\int_{-h}^{h} f(x)\,dx &\approx \frac{1}{2h} \cdot 2h - 0 + \left(\frac{1}{2}c^2\left(\Lambda'(a_i^m)\right)^2 - \frac{1}{2}c\Lambda''(a_i^m)\right) \cdot \frac{1}{2h} \cdot \frac{2}{3}h^3,\\
&= 1 + \left(\frac{1}{2}c^2\left(\Lambda'(a_i^m)\right)^2 - \frac{1}{2}c\Lambda''(a_i^m)\right) \cdot \frac{h^2}{3}.
\end{aligned}$$

$$\begin{aligned}
X_{\text{bin}}(a_i^m, h \mid \mu, \sigma, c) &\approx 1 - \exp\left(-c\Lambda(a_i^m)\right) \cdot \frac{1}{2h}\int_{-h}^{h} f(x)\,dx,\\
&\approx 1 - \exp\left(-c\Lambda(a_i^m)\right) \cdot \left[1 + \left(\frac{1}{2}c^2\left(\Lambda'(a_i^m)\right)^2 - \frac{1}{2}c\Lambda''(a_i^m)\right) \cdot \frac{h^2}{3}\right],\\
&= \underbrace{\left[1 - e^{-c\Lambda(a_i^m)}\right]}_{=X_{\text{mid}}(a_i^m \mid \lambda_a)} + e^{-c\Lambda(a_i^m)} \cdot \frac{h^2}{6}\left[c\Lambda''(a_i^m) - c^2\left(\Lambda'(a_i^m)\right)^2\right].
\end{aligned}$$

Thus, even with identical FOI parameters, the midpoint and binned seropositivity differ. This forward-process bias grows quadratically in the bin width $h$, and its sign depends on the local curvature of the FOI through $\Lambda'$ and $\Lambda''$:

$$X_{\text{bin}}(a_i^m, h \mid \mu, \sigma, c) \approx X_{\text{mid}}(a_i^m \mid \mu, \sigma, c) + \underbrace{\frac{h^2}{6} \cdot e^{-c\Lambda(a_i^m)} \cdot \left[c\Lambda''(a_i^m) - c^2(\Lambda'(a_i^m))^2\right]}_{\textbf{forward-process bias}},$$

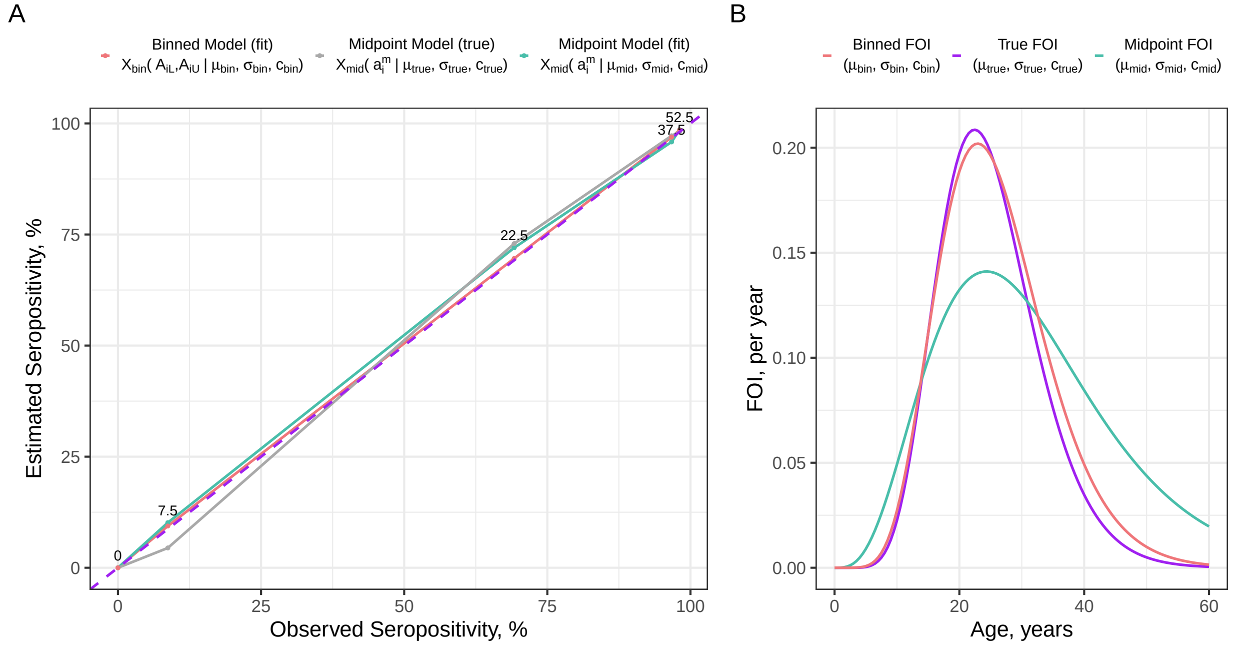


Figure S7: **Illustration of Forward-process and Inverse-process Biases in the Age-dependent FOI Model.** The underlying true FOI parameters used to generate data are $\mu = 25, \sigma = 8, c = 4$. Ages are grouped into 15-year age bins. **Panel A:** Observed versus estimated seropositivity at the bin level (one value per age bin, joined by lines for visual clarity). The legend indicates the seropositivity function representations used to calculate each seropositivity curve, as defined in Eq. S21 and Eq. (S20). The numbers shown next to the curves indicate the midpoint age (in years) that corresponds to each age bin. The observed seropositivities on the x-axis represent the proportion seropositive among individuals within each age bin in the data. The purple dashed line indicates perfect agreement, where the estimated seropositivity equals the observed value. The gray line shows the seropositivity evaluated at the midpoint age of each bin under the true age-dependent FOI. The green and red lines show the posterior predictive seropositivity obtained from the midpoint and binned model estimates, respectively. The gap between the diagonal purple line and the gray line represents the forward-process bias, and the shift from the gray to green line represents the inverse-process (inference) bias when the midpoint model adjusts its parameters to match the binned data. **Panel B:** Age-dependent FOI values that are supplied to Eq. S21 and Eq. S20 to calculate the corresponding seropositivities shown in Panel A: the true FOI (purple), the midpoint model FOI estimate (green), and the binned model FOI estimate (red).

Since $c^2(\Lambda'(a_i^m))^2$ is always positive, the sign of the forward-process bias is determined entirely by the second derivative of the cumulative exposure, $c\,\Lambda''(a_i^m)$. Under a gamma-shaped FOI, infection risk increases up to the peak age (around $\mu$) and declines afterwards. This means the corresponding gamma CDF has a second derivative that is positive before the peak, zero at the peak, and negative beyond it. Consequently, the sign of the forward-process bias is undetermined for younger ages (because both the first and second order derivatives are positive), but immediately after the peak age, we know that $\Lambda''(a_i^m) < 0$, which makes the forward-process bias negative. This implies that, for individuals older than the peak age, the midpoint model yields a seropositivity that is higher than the observed seropositivity. This comparison is illustrated in Figure S7A, where the gray line derived by supplying the true underlying FOI parameters into the midpoint model always lies above the diagonal purple dashed line for ages greater than $\mu = 25$.

##### S3.2.2 Inverse-process (Inference) Bias

When we fit the midpoint model to simulated datasets, for each age bin $k$, the data only provide the count of seropositive individuals $\#y_k$ out of $\#n_k$ individuals. Because we assume that individuals' exact ages within each bin are approximately uniformly distributed over $[a_k^m - h, a_k^m + h]$, the empirical seropositivity $\frac{\#y_k}{\#n_k}$ is an unbiased estimate of the bin-averaged seropositivity:

$$E\left[\frac{\#y_k}{\#n_k}\right] = \frac{1}{2h}\int_{a_k^m-h}^{a_k^m+h}\left[1 - e^{-c\Lambda(a)}\right] da = X_{\text{bin}}(a_k^m, h).$$

This highlights a key point: although we simulated data using the true underlying FOI, once ages are grouped into age bins, the observed datasets behave as if they were generated from the binned seropositivity curve (defined as the observed seropositivity on the x-axis in Figure S7A). However, from the forward-process bias, we know that:

$$X_{\text{bin}}(A_{iL}, A_{iU} \mid \mu, \sigma, c) = X_{\text{mid}}(a_i^m \mid \mu, \sigma, c) + \text{ forward-process bias},$$

because the midpoint model approximates each bin by evaluating seropositivity only at the midpoint age. As a result, the likelihood obtained from this midpoint approximation does not match the binned data (the gray line diverges from the purple diagonal line in Figure S7A), and this mismatch forces the model to compensate during inference.

To maximize the likelihood, the midpoint model adjusts its FOI parameters so that its posterior seropositivity (green) moves closer to the diagonal line. Under the gamma parametrization, this tends to push the model estimates toward a wider and lower FOI curve (i.e., larger $\sigma$ and a reduced peak). This adjustment is shown in Figure S7B, where the fitted midpoint FOI (green) becomes more spread out than both the true FOI (purple) and the binned FOI (red). In contrast, the FOI estimated by the binned model (red) closely aligns with the true underlying FOI (purple), as it correctly accounts for the age binning uncertainty. As expected, the posterior seropositivity curve evaluated using the binned model estimates is almost indistinguishable from the diagonal.

The biases described above, and illustrated for one specific parameter combination in Figure S7, can be generalized to other combinations of $(\mu, \sigma, c)$. A more comprehensive pattern can be visualized in Figure S8: across multiple parameter combinations, the midpoint model tends to infer FOI curves that are more spread out relative to both the true FOI and the binned model estimates and becomes more significant as the age bin size increases.

#### S3.3 Sensitivity Analysis of the Age-dependent FOI Model

For each parameter combination, we generate multiple datasets and repeated the inference several times. The plots shown here present representative results, which compare the estimated FOI from the exact, binned, and midpoint models against the true FOI curve used to generate the data, with shaded ribbons representing 95% credible intervals. Each row and each column corresponds to a different value of the scaling constant $c$ and age bin size used in model fitting. The parameters $\mu$ and $\sigma$ together define the age profile of infection risk, indicating whether the disease primarily affects childhood, adulthood, or elderly age groups. The scaling constant $c$ controls the overall FOI intensity within each scenario and is selected so that the peak FOI approximately corresponds to values of 0.05 to 0.1 and 0.2.

We use a one-at-a-time (OAT) sensitivity analysis framework, where we vary one parameter while holding others fixed to assess its individual effect. For each row in Figure S8, we vary $c$ to examine how the models behave as the peak FOI increases from approximately 0.05 to 0.1 and 0.2. From Figure S8(a) to Figure S8(b), the value of $\mu$ increases from 15 to 25 under a relatively sharp FOI profile (small $\sigma = 8$), while Figure S8(c) and Figure S8(d) assess the same trend under a more dispersed FOI profile (larger $\sigma = 20$).

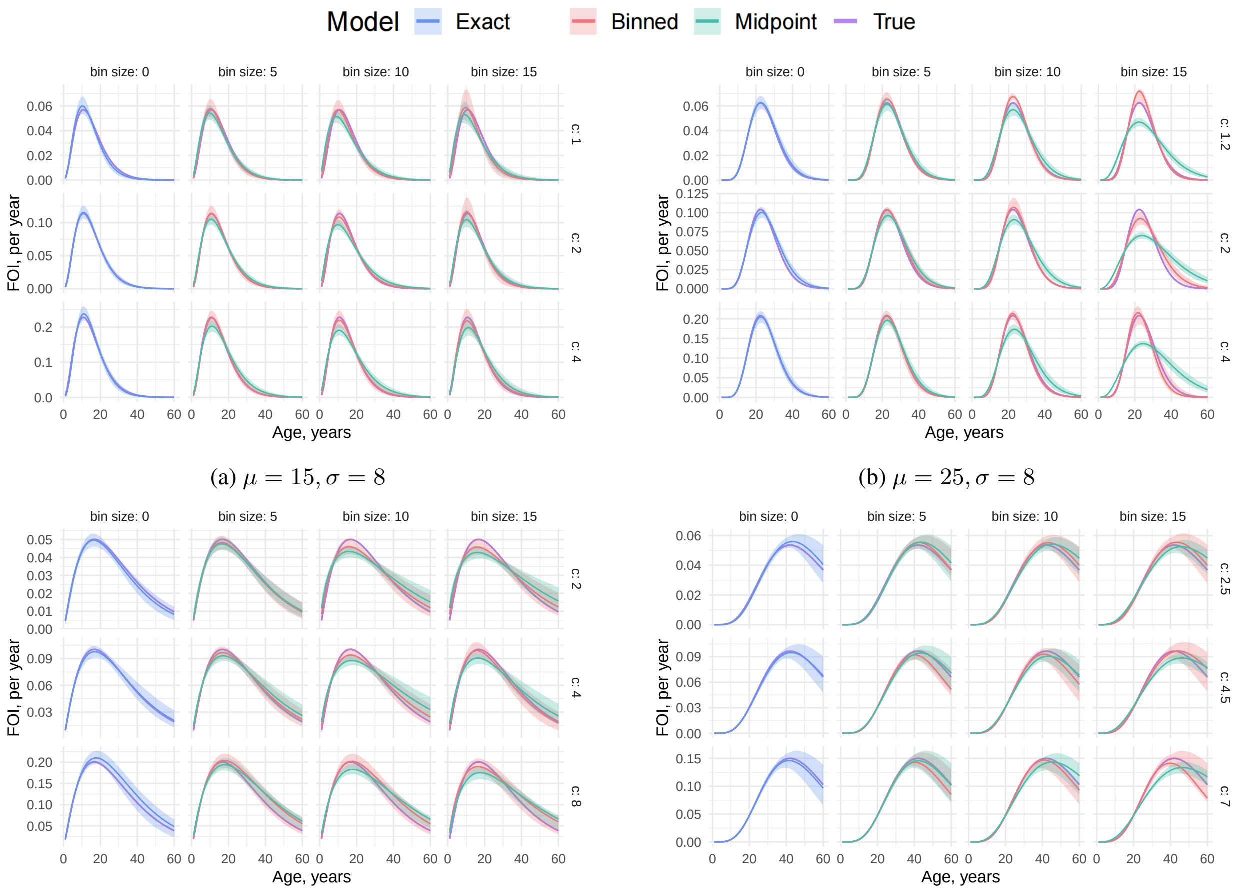


(a) $\mu = 15, \sigma = 8$

(b) $\mu = 25, \sigma = 8$

(c) $\mu = 30, \sigma = 20$

(d) $\mu = 50, \sigma = 20$

Figure S8: **Sensitivity analysis of the Age-dependent FOI Model.** Estimated age-specific FOI under varying parameter combinations of the gamma distribution $(\mu, \sigma)$ and scaling factor $c$. Each panel compares the Exact, Binned, and Midpoint models against the True FOI (purple). Columns correspond to different bin sizes (0, 5, 10, 15 years), and rows represent different values of $c$, chosen such that the true FOI peaks around 0.05, 0.1, and 0.2 per row within each panel.

#### S3.4 Sensitivity Analysis of the Time-dependent FOI Model

Figure S9 presents the estimated time-dependent FOI under a range of transmission patterns, including single peak epidemics, multiple waves, evasion in which FOIs progressively decrease over time (or increase over age), and endemic settings. This set of figures assumes that the FOI is piecewise-constant within 10-year intervals for both data generation and model fitting. In each plot, rows represent different scaling constants (i.e., transmission intensity), while columns correspond to different age bin sizes used to categorize individuals in a serosurvey. The true FOI (purple dashed) is compared against estimates from the exact model (blue), binned model (red), and midpoint model (green), with shaded ribbons indicating 95% credible intervals.

When age bin sizes are wider than the time period over which FOI is assumed constant (e.g., 20-year age bins vs. 10-year FOI intervals, shown as the rightmost panels in Figure S9(a)-(d)), inference becomes unreliable. Individuals within the same age bin may be exposed to multiple FOI levels, but the model cannot distinguish which parts of the bin correspond to changes in transmission. As a result, the likelihood provides weak information about when FOI shifts occur, making it difficult to recover changes in transmission. Indeed, the model shows clear convergence issues under these settings, with high $\hat{R}$ values and thousands of divergent transitions, indicating that the estimations are unreliable. Therefore, when individual ages are grouped into wide age bins in the serosurvey, it is important to consider whether the piecewise-constant FOI assumption remains reasonable for the level of resolution available in the data.

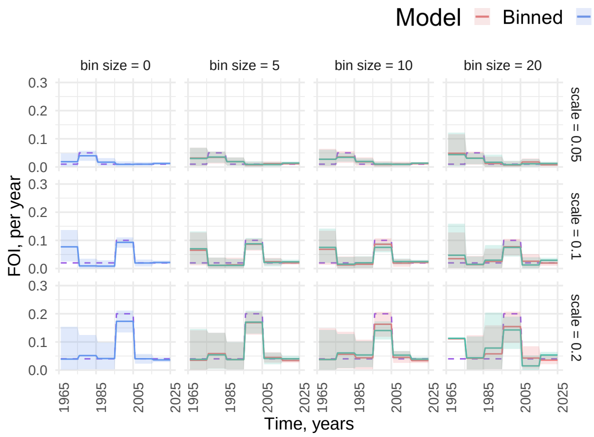


(a) Single Epidemic.

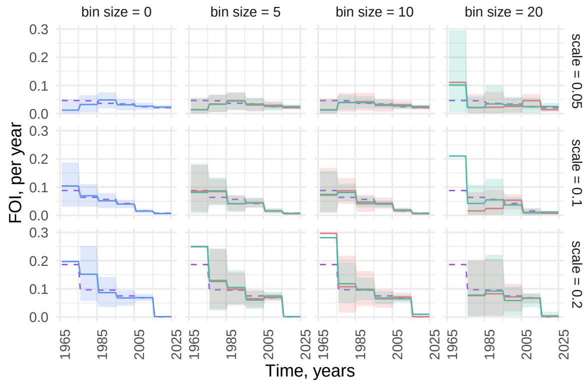


(b) Declining Transmission.

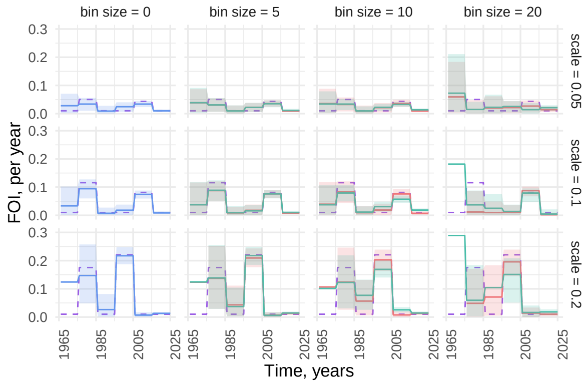


(c) Multiple Waves (Double Epidemics).

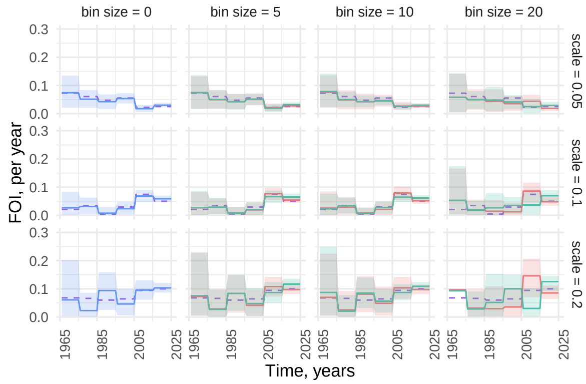


(d) Endemic.

Figure S9: **Comparison of FOI Estimation Scenarios Under 10-year intervals.** The $y$-axis is truncated at 0.3. **Panel (a)** Single Epidemic. The scale for each row controls the magnitude of a single FOI spike added to a constant low background FOI of 0.01 per year. **Panel (b)** Declining transmission. The scale represents the maximum possible FOI value that could be drawn from a uniform distribution. **Panel (c)** Multiple waves. A low background FOI of 0.01 per year is assumed throughout, and two random spikes are added: one in the earlier half of the age range, and one in the latter. The scale determines the magnitude of these spikes. **Panel (d)** Endemic. Infection risk changes with time, simulated using a random walk. The scale defines the maximum possible first FOI and determines how quickly FOIs can change between adjacent age intervals.

In Figure S10, we vary the interval length of the underlying FOI to 5-year intervals, while keeping the interval assumed during model fitting fixed at 10 years. This mismatch is important to consider because in real-world applications, the

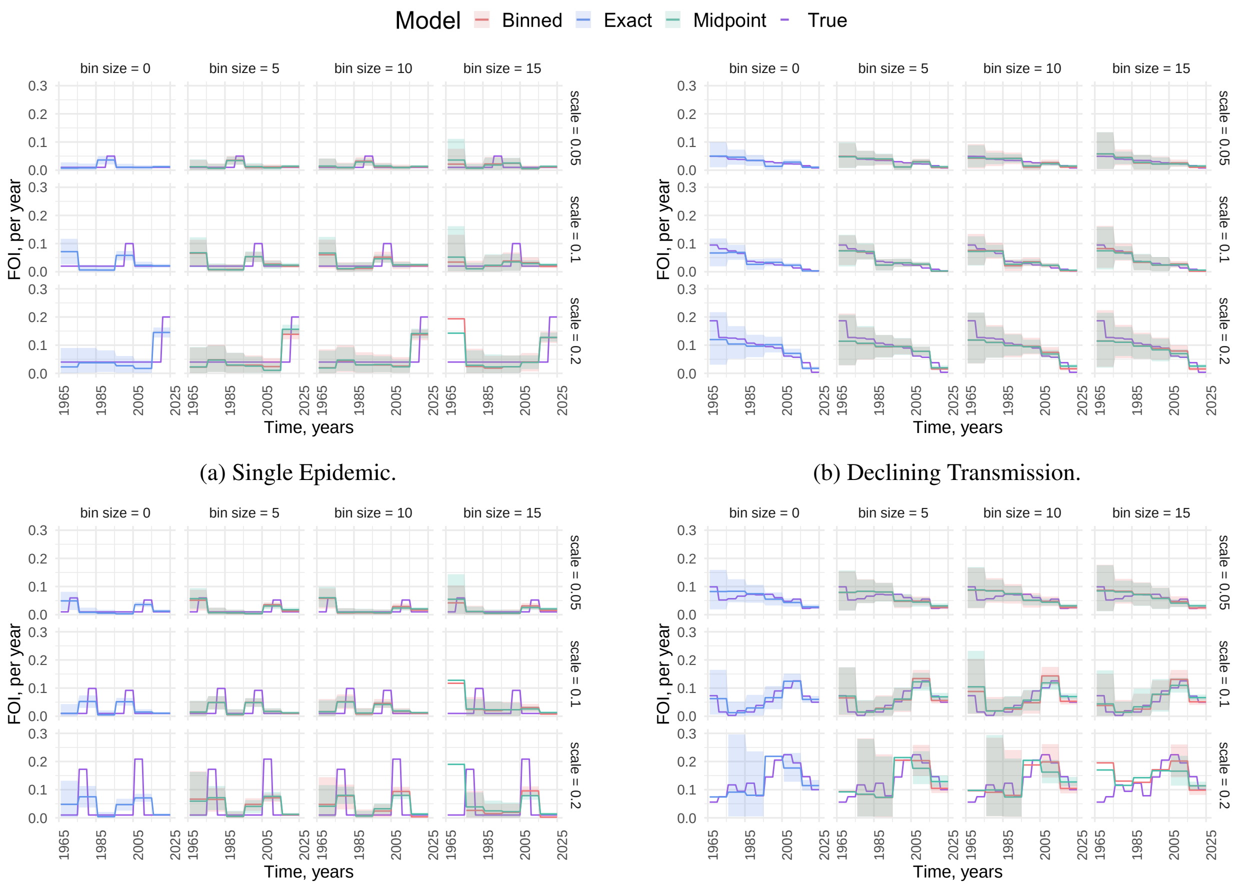


(a) Single Epidemic. (b) Declining Transmission.

(c) Multiple Waves. (d) Endemic.

Figure S10: **Comparison of FOI Estimation Scenarios under Interval Mismatch.** The $y$-axis is truncated at 0.3. In this set of simulations, the underlying FOI used to generate the data is piecewise-constant in 5-year intervals, whereas the model is fitted assuming 10-year constant intervals. Relative to the true FOI (purple, solid line for clearer visualization), estimates from the exact model (blue), binned model (red), and midpoint model (green) are shown, with shaded ribbons indicating 95% credible intervals. Same transmission patterns are presented as in Figure S9.

true intervals over which the FOI remains constant are unknown, and analysts must make an informed but subjective choice based on data availability and sample size.

Interestingly, even under this mismatch, the models are still able to capture the general temporal fluctuations in transmission. However, because the fitted intervals are wider, changes in FOI are spread over a longer time window, leading to lower estimated FOIs compared to the true FOI in the single- and double-epidemic scenarios, which is expected. In the evasion setting, the model performs reasonably well, with estimates roughly corresponding to averages across neighboring 5-year intervals. For the endemic case, the models continue to recover the overall trend, but, as expected, performance worsens as the bin width increases.

### References


[1] W. M. Landau. "The targets R package: a dynamic Make-like function-oriented pipeline toolkit for reproducibility and high-performance computing". *Journal of Open Source Software* 6.57 (2021), p. 2959.

[2] R Core Team. *R: A Language and Environment for Statistical Computing*. R Foundation for Statistical Computing. 2024. URL: `https://www.R-project.org/`.

[3] Stan Development Team. *RStan: the R interface to Stan*. 2024. URL: `https://mc-stan.org/`.
